# Creating a computer assisted ICD coding system: performance metric choice and use of the ICD hierarchy

**DOI:** 10.1101/2024.01.16.24301382

**Authors:** Quentin Marcou, Laure Berti-Equille, Noël Novelli

## Abstract

**Objective:** Machine learning methods hold the promise of leveraging available data and generating higher-quality data while alleviating the data collection burden on healthcare professionals. International Classification of Diseases (ICD) diagnoses data, collected globally for billing and epidemiological purposes, represents a valuable source of structured information. However, ICD coding is a challenging task. While numerous previous studies reported promising results in automatic ICD classification, they often describe input data specific model architectures, that are heterogeneously evaluated with different performance metrics and ICD code subsets.

This study aims to explore the evaluation and construction of more effective Computer Assisted Coding (CAC) systems using generic approaches, focusing on the use of ICD hierarchy, medication data and a feed forward neural network architecture.

**Methods:** We conduct comprehensive experiments using the MIMIC-III clinical database, mapped to the OMOP data model. Our evaluations encompass various performance metrics, alongside investigations into multitask, hierarchical, and imbalanced learning for neural networks.

**Results:** We introduce a novel metric, RE@R, tailored to the ICD coding task, which offers interpretable insights for healthcare informatics practitioners, aiding them in assessing the quality of assisted coding systems. Our findings highlight that selectively cherry-picking ICD codes diminish retrieval performance without performance improvement over the selected subset. We show that optimizing for metrics such as NDCG and AUPRC outperforms traditional F1-based metrics in ranking performance. We observe that Neural Network training on different ICD levels simultaneously offers minor benefits for ranking and significant runtime gains. However, our models do not derive benefits from hierarchical or class imbalance correction techniques for ICD code retrieval.

**Conclusion:** This study offers valuable insights for researchers and healthcare practitioners interested in developing and evaluating CAC systems. Using a straightforward sequential neural network model, we confirm that medical prescriptions are a rich data source for CAC systems, providing competitive retrieval capabilities for a fraction of the computational load compared to text-based models. Our study underscores the importance of metric selection and challenges existing practices related to ICD code sub-setting for model training and evaluation.

Statement of significance

Problem or Issue
Accurate ICD coding is challenging and time consuming, leading to low data quality.

What is Already Known
Machine learning algorithms, could improve ICD coding but their use in production environments remains limited. Existing work focuses on input data specificities and automated classification on heterogeneous subsets of ICD codes.

What this Paper Adds
We introduce interpretable performance metrics tailored for computer assisted coding, and identify target metrics to improve ranking performance. We show that utilizing the full set of ICD codes is beneficial even for input data with seemingly low information. Furthermore, we explore multitask, hierarchical and class imbalance correction methods demonstrating their limited benefits.

## I. Introduction

The different versions of the International Classification of Diseases (ICD) have been used for annotating clinical data in tens of countries for several decades. While this annotation is primarily used for healthcare billing and planning purposes, it should also constitute a wealth of structured data for large-scale epidemiological studies and personalized predictions [1].

However, accurate ICD coding is both difficult and time consuming. This coding difficulty results in rather low data quality, e.g., with inter-coder agreement between fair and poor for principal diagnostic coding at the billing level, even for professional coders [2].

With the release of freely-available datasets such as MIMIC [3], there has been an endeavor of computer science and health informatics researchers to help the medical community with this clinical coding burden. In the last decade, many different approaches from rule-based algorithms [4] to supervised learning algorithms (such as Support Vector Machines (SVMs) [5, 6] or Neural Networks (NNs)) have been explored. The NN approach is the most prevalent nowadays. NNs are particularly used for automated ICD coding based on unstructured text (using Transformers [7–9], CNNs [10– 15] or RNNs [16–18]) but also more marginally for ICD automated coding based on structured [19, 20] or multimodal [11, 21] data (see Table S1 for a quick review of existing methods on the MIMIC-III database). The utilization of medication information in ICD coding has been relatively understudied with, to our knowledge, only one published study on this topic [20]. Nevertheless, medication information may present several assets: 1) It is closely linked to actual diagnosis for medical conditions; 2) Systematic recording by prescription softwares; 3) It is language-insensitive and uses normalized nomenclatures such as RxNorm; 4) Anonymization for multi-center training is straightforward; 5) Medical treatments follow strict recommendations, offering a priori good generalizability; 6) Being represented as structured data, decent performance could be expected even with a simple model and limited preprocessing.

Despite promising results, a limited number of studies document the use of such methods in a production environment [22, 23]. The adoption of such systems by the medical informatics community may be hindered by several factors. For one, conscious of the limited quality of training data and the difficulty of the coding task, coders seem to expect Computer Assisted Coding (CAC) systems, i.e., semi-automatic systems with human in the loop systems, rather than fully automated coding systems [24, 25]. In turn, commonly reported performance metrics such as F1 score and analysis on small code subsets, such as top 50 billing codes (see Table S1), may seem irrelevant for the clinical coding community. Finally, while existing studies concentrate on optimizing input-specific model architectures, a notable gap lies in the underexplored territory of leveraging the inherent properties of the ICD hierarchy, a feature inherent to any ICD coding system and model architecture. Indeed, albeit few studies using NNs leverage the ICD hierarchy information using an attention mechanism in conjunction with a graph NN [14, 15, 26, 27] or text labels [7, 11, 16] within their architecture, only SVMs based studies [5, 28] have made use of generic, model architecture agnostic, Hierarchical Multilabel Classification (HMC) techniques. Similarly, class imbalance, though inherent to HMC, is also seldom addressed.

In this paper, we present several significant model architecture-agnostic contributions to ICD coding using a CAC system through a systematic study of techniques to exploit CAC systems and ICD properties. First, we define a set of performance metrics, RE@R, tailored to the ICD coding task, and ensuring interpretability by clinical coders who may not be experts in machine learning. Second, we investigate and compare various performance metrics, revealing that optimizing NDCG or precision-recall based metrics leads to improved ranking performance compared to traditional F1 based metrics. Third, we examine the impact of utilizing the entire set of ICD codes instead of cherry-picked subsets, finding that it maintains performance on selected codes and enables more efficient recovery of additional codes from seemingly limited medication input data. Fourth, we conduct a systematic study of generic approaches with low computational overhead to test whether exploiting the hierarchical properties of the ICD classification can improve CAC systems. This includes multitask learning scenarios, hierarchical multilabel classification techniques, and class imbalance correction techniques. Our findings suggest that NNs benefit from learning from the whole hierarchy; however, neither HMC nor class imbalance correction methods improved results upon the simple multitask learning NNs. Finally, our study serves as a valuable replication of predicting ICD-9 codes based on medication information [20] using the MIMIC-III dataset [3], achieving superior classification and ranking results, as well as promoting broader applicability through the use of the OMOP-CDM data-standard.

## II. Material and Methods

### A. Data

#### 1) MIMIC-III OMOP

We performed all our experiments on the MIMIC-III v1.4 clinical database [3], a freely-available database of 58,976 ICU stays. The raw database was mapped to the OMOP-OHDSI common data model using scripts from [29], in order to exploit the mapping of the non-standard drug prescription representation in MIMIC-III to the RxNorm standard.

#### 2) Medication prescription data

We then transformed the resulting RxNorm codes into their corresponding RxNorm ingredients, thereby discarding any clinical form, dose or brand information. This process resulted in a set of 1,164 unique RxNorm ingredients. Subsequently, we further simplified the medication prescription data by transforming it into one binary hot-encoded vector per stay, indicating whether a particular RxNorm ingredient had been prescribed at least once during the patient’s stay.

#### 3) ICD9-CM

MIMIC-III diagnoses use the ICD9CM nomenclature, which is hierarchical with a maximal depth of 5 levels denoted, in increasing depth, as Chapters, Subchapters, 3 digits, 4 digits, and 5 digits codes. Stays are generally annotated only by leaf nodes, as only leaf nodes are used for billing purposes. Notably, not all branches have the same depth, resulting in Billing codes of either 3, 4, or 5 digits.

In order to obtain complete annotations for each ICU stay, we performed a roll-up of the ICD9 hierarchy. The resulting numbers of unique codes and the numbers of codes per ICU stay are shown in Table I and Table II, respectively.

**TABLE I:** Number of unique codes per level. The cherry picked subset corresponds to a subset of codes filtered on both frequency and a prior belief about the potential amount of information contained in the input data, as defined in Section II-A4.

|  | ICD9-CM | MIMIC III | Cherry picked |
| --- | --- | --- | --- |
| Chapters | 19 | 19 | 16 |
| Subchapters | 154 | 151 | 68 |
| 3 digits | 1,234 | 1,070 | 312 |
| 4 digits | 7,473 | 4,380 | 520 |
| 5 digits | 8,846 | 3,716 | 318 |
| Billing | 12,167 | 5,871 | 613 |
| All levels | 17,726 | 9,336 | 1,234 |

**TABLE II:**
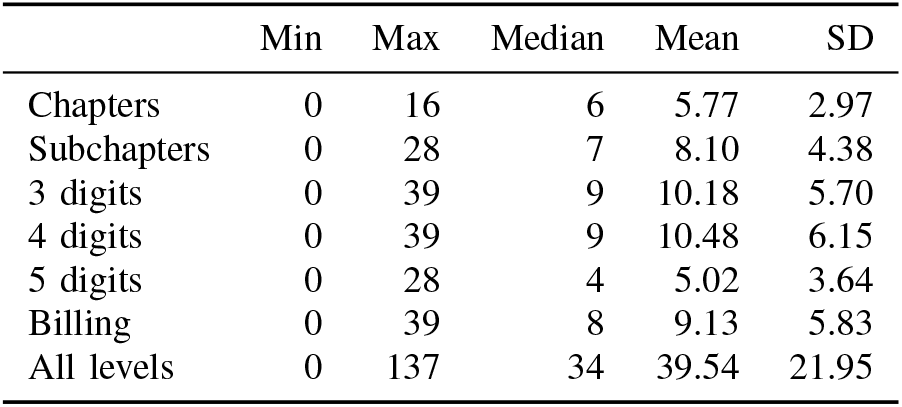
Number of codes per stay for different ICD levels on the complete MIMIC-III code set.

|  | Min | Max | Median | Mean | SD |
| --- | --- | --- | --- | --- | --- |
| Chapters | 0 | 16 | 6 | 5.77 | 2.97 |
| Subchapters | 0 | 28 | 7 | 8.10 | 4.38 |
| 3 digits | 0 | 39 | 9 | 10.18 | 5.70 |
| 4 digits | 0 | 39 | 9 | 10.48 | 6.15 |
| 5 digits | 0 | 28 | 4 | 5.02 | 3.64 |
| Billing | 0 | 39 | 8 | 9.13 | 5.83 |
| All levels | 0 | 137 | 34 | 39.54 | 21.95 |

#### 4) Cherry-picked ICD9 codes

As shown in Table S1 many studies investigating automatic ICD coding using MIMIC-III focus on a cherry-picked subset of ICD9 codes. In particular in Hansen et al. [20], the only existing study focusing on the use of medication to predict ICD diagnoses, codes were cherry-picked according to both frequency and researchers’ prior belief regarding medication’s information content about the different diagnoses. Aiming to reproduce Hansen et al. [20] ‘s filters and construct our cherry-picked code set, we discarded the following codes:

- codes with less than 100 occurrences in the complete dataset;
- codes belonging to chapters *Injury And Poisoning* (800-999), *Supplementary Classification Of External Causes Of Injury And Poisoning* (E000-E999), and *Supplementary Classification Of Factors Influencing Health Status And Contact With Health Services* (V01-V91);
- codes belonging to *Disorders relating to short gestation and low birth-weight* (765 3-digits code).

The resulting number of unique codes and number of codes per ICU stay for the cherry-picked code set are respectively shown in Table I and Table S2. Note that despite our efforts to replicate their code filtering we ended up retaining significantly more codes than reported in Hansen et al. [20].

### B. Models

#### 1) Architecture

As the emphasis of our study is not to optimize a model structure specific to drug prescriptions, but rather perform a systematic evaluation of techniques making use of the ICD hierarchy properties, we favored a computationally frugal approach and used simple sequential NNs with hyperparameters described in Table S3.

Our neural networks take as input a vector of length 1,166 containing:

- a binary indicator for patient’s sex;
- the age of the patient clipped and normalized by 89 years, the maximum age reported in MIMIC;
- 1,164 binary indicators for hot encoded RxNorm ingredients prescribed at least once during the patient’s stay.

Unless specified otherwise, we use a sigmoid activation for the last dense layer. During training, we implement an early-stopping strategy by monitoring the target performance metric evaluated on 5% of the training set used as a development validation set. All models were implemented using Tensorflow 2.9.1. We performed hyperparameter optimization using 2 passes of the Hyperband algorithm [30], an early-stopping based hyperparameter tuning strategy implemented in the Keras-Tuner package.

We performed a three-fold Monte Carlo crossvalidation. For each fold, we sampled 5,000 random visits without replacement for the test set, 5,000 for the development set, and used the remaining 48,976 visits as the training set. The same three cross-validation sets were used for the different experiments. We used paired statistical tests accordingly when necessary.

#### 2) Dummy model

For the sake of comparison, we introduce a dummy ranking model that ranks codes solely based on their frequencies estimated from the training set. Consequently, the predictions of this dummy model remain constant for any input.

### C. Performance metrics

Table S1 presents research conducted on ICD code prediction using the MIMIC-III dataset, where various performance metrics, primarily derived from the field of automatic classification, have been employed. However, these metrics might not be ideal for evaluating a recommendation system designed to assist clinical coders in the coding process. Their practical significance for clinical coders, who are the primary users, could be particularly challenging to grasp. In turn, this lack of clarity may hinder the adoption of CAC systems. In this section, we introduce a dedicated set of metrics of performance quantifying ranking errors. We defer the definition of other usual metrics that we will use as comparison to Appendix C.

Our objective is to identify a performance metric that possesses the following essential characteristics:

- it captures the idea that all relevant codes must be retrieved by the clinical coder;
- it quantifies each non-relevant code mistakenly ranked before a relevant one as one unit of lost time for clinical coders;
- it allows for meaningful comparisons between different datasets or hospital wards;
- it can be easily understood and interpreted by clinical coders and healthcare informatics staff who may not have expertise in machine learning.

While widely used for assessing the performance of recommender systems, Normalized Discounted Cumulative Gain (NDCG) presents challenges in interpretation due to its nonlinear discounting, which assigns different weights to errors based on label rank. Additionally, its value is influenced by the number of labels considered [31], making it difficult to compare models trained on different code subsets or databases. In contrast, Precision@Recall implicitly assigns different weights to errors based on the number of positive labels per sample. Recall@K is more straightforward to interpret, but the choice of the relevant K depends on the average number of codes expected to be found in a given patient stay. For example, a clinical coder may be willing to a longer list of recommended codes annotating an ICU stay than annotating a simple day clinic visit for chemotherapy. Lastly, `Coverage`, a metric that measures the number of labels that must be examined in a ranked list to achieve 100% recall, is easily interpretable but also influenced by the number of positive labels per stay.

We introduce a set of related metrics RankErrors@Recall or RE@R. These metrics represent the number of negative labels that are mistakenly ranked above the positive labels until a fraction *R* of the positive labels is recovered. This set of metrics is a generalization of the notion of coverage made independent of the number of positive labels per example:

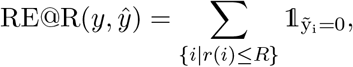

where *r*(*k*) is the recall at rank *k*, or Recall@K, and 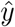 the vector of ground truth labels sorted in decreasing order according to the predicted score *ŷ*. Note that given this definition:

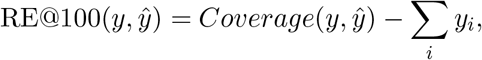

meaning that the sample averaged RE@R is simply a more granular version of coverage that is independent of the average number of positive labels per sample in the dataset.

### D. Multitask learning

Multitask Learning (MTL) is the process of training an algorithm to perform multiple related tasks simultaneously, with the aim to improve both performance and generalization compared to isolated task training [32]. In that sense, performing multilabel ranking or classification for a single level of the ICD hierarchy, already is an instance of multitask learning.

In the remaining of this paper, we will however refer to the notion of multitask with the hierarchical nature of the ICD in mind, where ranking or classification at each level of the hierarchy corresponds to a different task. We evaluate whether learning simultaneously on different hierarchy levels is beneficial using three distinct learning strategies:

- Learning per level: a different model is trained for each level of interest in the hierarchy (Chapters, Subchapters, 3-digits and Billing levels);
- Naive multitask learning: a flat classifier is trained with an output layer containing an output neuron for each node of the hierarchy (9,336 labels);
- Re-weighted multitask learning: by construction, there exist more labels at each level as we progress towards the leaves of the hierarchy, resulting in higher importance of finer grained levels of the hierarchy in the cost function. To give equal importance to each task, or level, we have re-weighted the cost of each example such that each task have the same weight in the cost function (Appendix E3).

### E. Hierarchical Multilabel Classification (HMC)

While multitask learning leverages the ICD hierarchy in an implicit manner, dedicated methods exist to explicitly exploit the known hierarchical links in multilabel classification or ranking. In our study, we implemented several of them, employing different approaches to incorporate hierarchical knowledge. These include the use of simple logical rules (roll-down and roll-up) to enforce hierarchical consistency of predictions, modifications of the cost functions based on logical rules such as TreeMin loss [33] and MCLoss [34], as well as hierarchical regularization techniques [35] (Appendix F).

### F. Class imbalance

Large scale hierarchical problems inherently exhibit imbalanced label distributions, often following a powerlaw like distributions for deep hierarchies [36] (Fig. S1). Severe imbalance can lead an algorithm to ignore positive examples of the minority class to limit the amount of false negative for the majority class, or lead to poor generalization on a dataset with different class frequencies. In classification settings, class imbalance can be mitigated using regularization, cost function modifications or more frequently using resampling methods. Despite being the most frequent, resampling approaches increase computational load and impose constraints on model types and learning objectives for HMC. Thus, we focused on applying a regularization approach, using L2 regularization on the last classification layer, and three cost sensitive approaches: two based on upweighting the cost of positive examples by the imbalance ratio, namely Imbalance Ratio Weighting (IRW) and Imbalance Ratio Normalized Weighting (IRNW), and one variant of the cross-entropy loss, initially used in computer vision, and referred to as focal loss (Appendix G).

## III. Results

### A. ICD code cherry picking degrades retrieval performance

A distinctive aspect of medication data as input for building a CAC system is that medication could be expected to convey little to no information about some diagnoses, such as, for example, diagnosis codes belonging to the 3-digits node *V50-V59 Persons Encountering Health Services For Specific Procedures And Aftercare*. This has led some authors to design algorithms only on cherry-picked subsets of codes filtered both by code frequency and researchers’ prior beliefs about the information content of input data regarding ICD codes [20] (see Section II-A4). Such filtering can be justified assuming that 1) the input data conveys no information about some diagnostics and/or 2) that data is too scarce to allow any learning and/or 3) that the inclusion of such codes in the model deteriorates predictions for the selected cherry-picked codes.

We challenged the validity of these first two assumptions by training models on the complete code set and quantifying the amount of information a model, using a naive multitask architecture and trained to maximize *μ*F1, extracted for different code subsets. Figure 1a shows the resulting cumulative distribution of per-code entropy reduction, i.e., the reduction in uncertainty regarding the presence or absence of a code once the model’s output is known. We find that, despite a generally lower information content, the model was still able to extract some information for more than 75% of low frequency codes with ≤100 occurrences in the entire MIMIC dataset, with a reduction of entropy of at least 10% for 31.8% of such codes. In fact, the algorithm was able to extract some information even for some codes with less than 5 occurrences in the whole dataset (see Fig. S2). Strikingly, Figure 1a also suggests that codes filtered out based on researcher’s prior beliefs about information content are not harder to predict than cherry-picked codes. We performed a Mann-Whitney U test and found that the entropy reduction distribution was comparable for both code sets (p=.155).

**Fig. 1:**
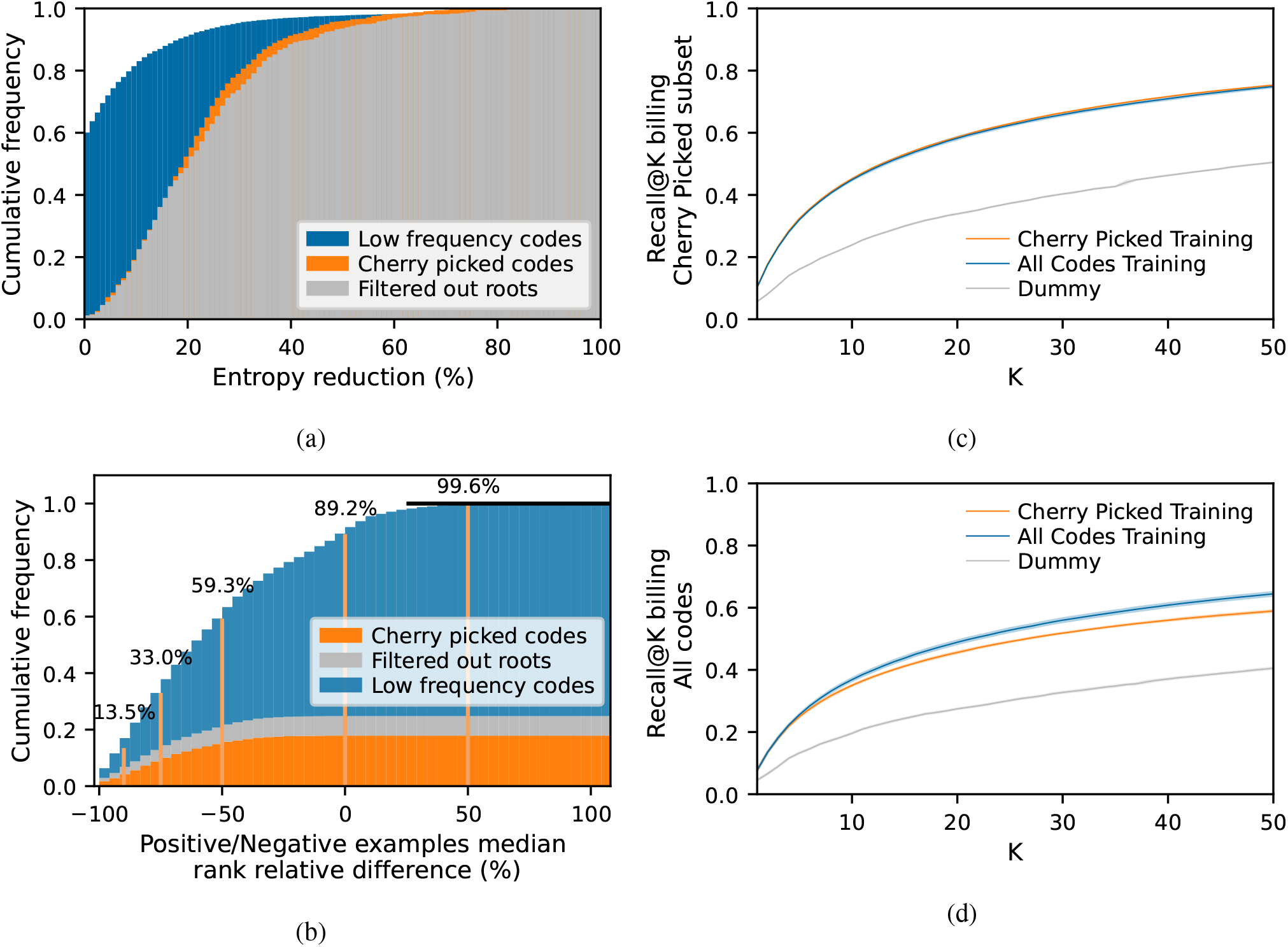
**a**. Cumulative distribution of the entropy reduction, or information gain, per ICD code at any level of the hierarchy by performing soft classification using our naive multitask neural network model. Low frequency codes designate labels with ≤ 100 occurrences in the complete dataset. Filtered out roots designate codes excluded based prior beliefs (Section II-A4) **b**. Cumulative distribution of the relative difference of median ranks between positive and negative examples for each ICD code. ICD codes that are, as expected, better ranked on positive than on negative examples exhibit negative values, while codes with worse ranking on positive examples have positive values. For legibility, we added a horizontal black line at y=1. **c**. Recall@K on the cherry picked code subset using a dummy model (green), or neural networks models trained respectively on the cherry picked subset only (blue) or the complete code subset (orange). Shades around the lines show the standard error on the estimated sample averaged Recall@K computed via three-fold Monte-Carlo cross-validation. **d**. Recall@K on the complete code set using the same models. ICD codes not seen at training time by the model trained on the cherry picked subset only were assigned 0 probability at inference time. The four sub-figures were made using models built to optimize *μ*F1.

Figure 1b illustrates how this extracted information content translates into ranking capabilities by comparing ranks for positive examples versus ranks for negative examples for a given code. 89.2% of codes were better ranked for positive examples than negative examples, 59.3% of codes had their rank at least halved on positive examples and 13.5% had their rank on positive examples being less than a tenth of their rank on negative examples. Every code in the cherry-picked subsets or filtered out codes based on researcher’s prior beliefs had better ranks on positive examples, and 83.65% of them had a rank more than halved on positive examples.

Then, we challenged the assumption that adding codes on which the algorithm would fail to extract information could worsen the algorithm’s predictions on cherrypicked codes. To that end, we carried hyperparameter optimization and training both using the complete code set and the cherry-picked code subset. Fig. 1c, displaying the Recall@K on the cherry-picked code subset, shows that both models had equivalent code retrieval performance on that subset. Similar conclusions could be drawn for automatic coding systems, relying on hard classification as shown by the *μ*F1 scores in Table S4.

Finally, one might still wonder whether cherry-picking codes actually matters from a clinical coder perspective. Fig. 1d shows that using a model trained on the complete code set does improve Recall@K on billing codes even for `K` as low as 9, the average number of billing codes per stay (see Table II), with for instance 4.16% of added recall for `K`=30.

### B. NDCG and AUPRC variants are better target metrics than F1

ICD codes prediction can be viewed as two different tasks: fully automatic coding or building a coding assistant system. The latter can be seen as a learning to rank problem with the specificity that all codes that must be coded should be found in the recommendations. While building a perfect classifier for automatic coding entails building a perfect recommender system, classical classification metrics may not convey complete or intuitive information on how useful an imperfect soft classification algorithm is as a coding assistant.

Having this hybrid system between multi-label classification and recommender or learning to rank system in mind, we defined the RE@R set of metrics in Section II-C. This metric denotes the average number of non relevant codes that have to be seen by a clinical coder browsing the ranked list of codes before achieving a recall of `R`.

While achieving the minimal RE@100 of 0 would mean achieving a perfect recommender system, RE@100 alone is insufficient to describe the performance of an imperfect recommender system, and RE@R for different values of `R` is of interest. Trying to minimize RE@R for different values of `R` at the same time would, however, be impractical. We thus searched for a more traditional scalar performance metric that by optimizing, we would also minimize RE@R for different values of `R`. With that aim, we performed hyperparameter search and training on the complete code set targeting various widely used ranking and classification metrics. The results are shown in Fig. 2, as relative RE@R improvement over a model selected and trained trying to optimize the *μ*F1 score.

**Fig. 2:**
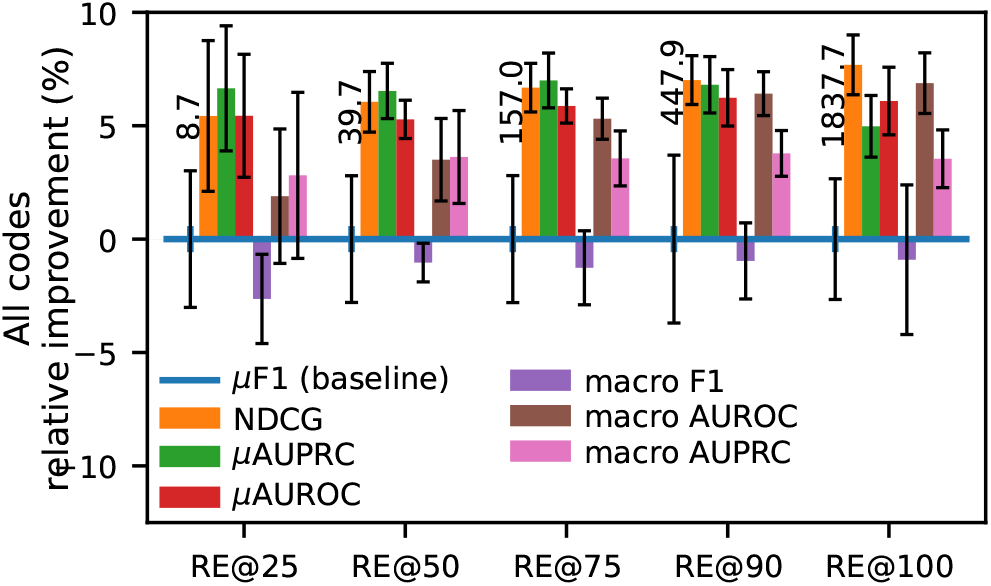
Relative improvement in ranking performance, measured by RE@R, for different values of R achieved by optimizing various scalar performance metrics instead of the commonly reported *μ*F1. Error bars represent the standard error of the mean computed over crossvalidation runs. We indicate, as reference, the absolute RE@R values, averaged over cross validation runs, achieved by the baseline model optimizing the *μ*F1.

Despite being by far the most reported performance metric for automated ICD coding on MIMIC (see Table S1), Fig. 2 shows that optimizing F1 based metrics leads to lower ranking performance, and that optimizing either NDCG or a micro averaged AUC variant leads to ranking performance improved by ∼5% for any recall. In general, macro-averaged metrics seem to lead to lower ranking performance compared to their micro averaging counterparts. Similar conclusions can be drawn on a per hierarchy level basis from Fig. S3. To further strengthen these points, without extra model training computations, we show in Fig. S4 taking into account all models we have trained, that NDCG has highest correlation with RE@R for all values of `R`, closely followed by Precision-Recall based metrics.

Unsurprisingly, the NDCG, a dedicated ranking metric, seemed to be the most effective target metric to optimize ranks. However, because the computation of NDCG only relies on rank, it may not always prioritize models whose predictions can be readily interpreted as probabilities. Interestingly, our best NDCG models already demonstrated strong calibration. Nevertheless, our models chosen to maximize Area Under the Precision Recall Curve (AUPRC) exhibited notably enhanced calibration, reflected in significantly improved Expected Calibration Error (ECE) and Maximum Calibration Error (MCE) values (Table S5).

### C. Multitask learning improves ranking and runtime

In this section, we investigate whether a naive use of the hierarchy information, by training a NN to rank (or classify) simultaneously codes from different levels of the hierarchy, can impact performance. Using multitask learning a NN may discover and exploit hierarchical relationships between tasks in hidden layers [32].

We employed three distinct training methodologies: per-level training, which yielded four separate models; a naive multitask approach; and a re-weighted multitask strategy, where equal weight was assigned to each hierarchy level in the loss function (see details in Section II-D). As depicted in Figure 3, when using NDCG as a ranking performance indicator, the re-weighted multitask approach led to a significant degradation in performance across all levels of the ICD hierarchy. In contrast, while the average NDCG per level was generally higher for the naive multitask approach compared to the per-level strategy, the observed differences remained within error bounds (refer to Figure 3). Similarly, looking at RE@R (Fig. S5), the estimated performance was greater for the naive multitask model across various values of R and all hierarchy levels when compared to the per level models. These estimated differences, performing a paired t-test, were however deemed statistically significant only at the Billing level and for recalls higher than 75%. Notably, hyperparameters selected for the per-level models closely resembled those obtained for the naive multitask model (see Table S10). This observation indicate a runtime advantage associated with the naive multitask approach, which scales linearly with the number of hierarchy levels.

**Fig. 3:**
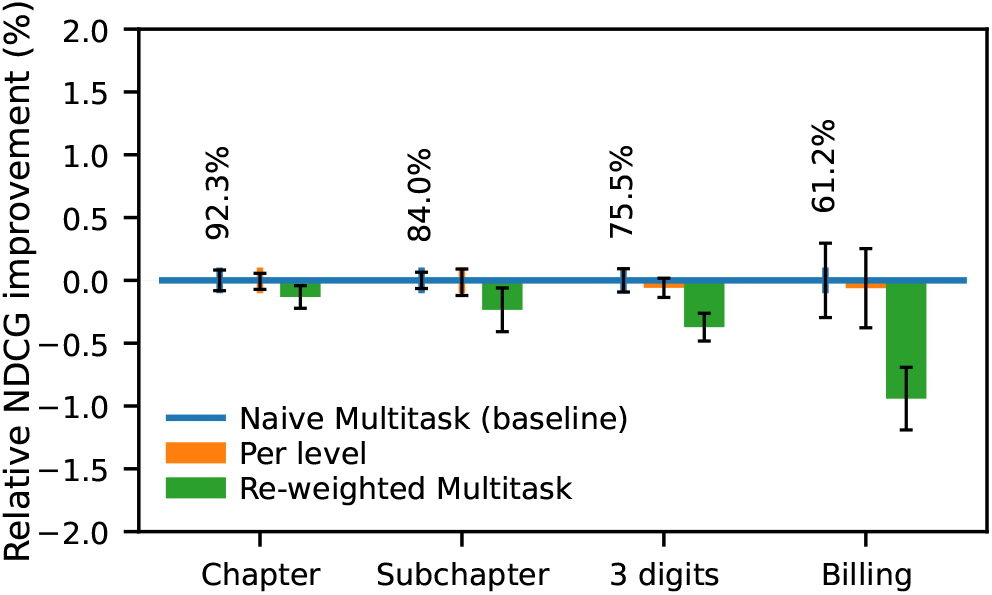
Relative improvement in ranking performance, measured by NDCG, across different levels of the ICD hierarchy using various multitask learning strategies compared to the baseline naive multitask model. Absolute NDCG values, averaged over cross-validation runs, for the baseline naive multitask model are provided as reference.

### D. Hierarchical learning methods do not improve ICD code retrieval

In the previous section, we have shown that simply adding hierarchy levels as complementary tasks only marginally improves performance. In this section, we investigate whether explicit use of known hierarchical relationships, at learning and/or inference time, can further improve performance. We compared a flat classifier (naive multitask) with a diverse set of global hierarchical approaches using logical constraints (roll-up and roll-down), special cost functions (MCLoss, TreeMin), or regularization (HierL2) (see Section II-E).

The NDCG achieved by these different models are illustrated in Fig. 4.We find that, analyzing performance based on NDCG, none of the hierarchical methods tested significantly change ranking performance. In fact judging by rank errors statistics, the two cost based methods, TreeMin and MCLoss, even seem to degrade ranking performance on billing codes below 50% recall (Fig. S5).

**Fig. 4:**
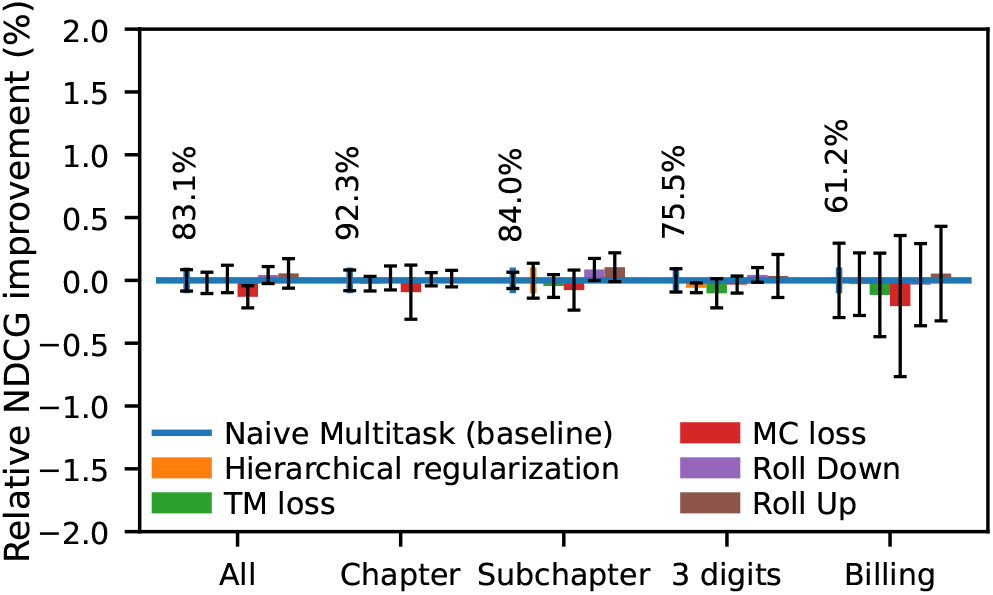
Relative improvement in ranking performance, measured by NDCG, across different levels of the ICD hierarchy using various hierarchical multilabel classification strategies compared to the baseline naive multitask model. Absolute NDCG values, averaged over crossvalidation runs, for the baseline naive multitask model are provided as reference.

### E. Class imbalance correction does not benefit ICD codes retrieval

As illustrated in Fig. S1, the breadth and depth of the ICD classification result in highly imbalanced labels. Taking into account class imbalance has proven beneficial for some classification tasks, but has been less studied for recommender systems.

We have implemented and applied several cost sensitive and regularization based methods used for imbalanced classification (see Section II-F) to assess whether addressing class imbalance could improve our recommending system. Our results, shown in Fig. 5, suggest that these different methods either did not improve or even worsened the global ranking performance at every level of the ICD hierarchy. In fact, our baseline naive multitask model seem to exploit jointly code frequency (Fig. S6) and input data information (Fig. 1) for rank predictions, while still being able to output high ranks (e.g., top-10) for some very low frequency codes (Fig. S6).

**Fig. 5:**
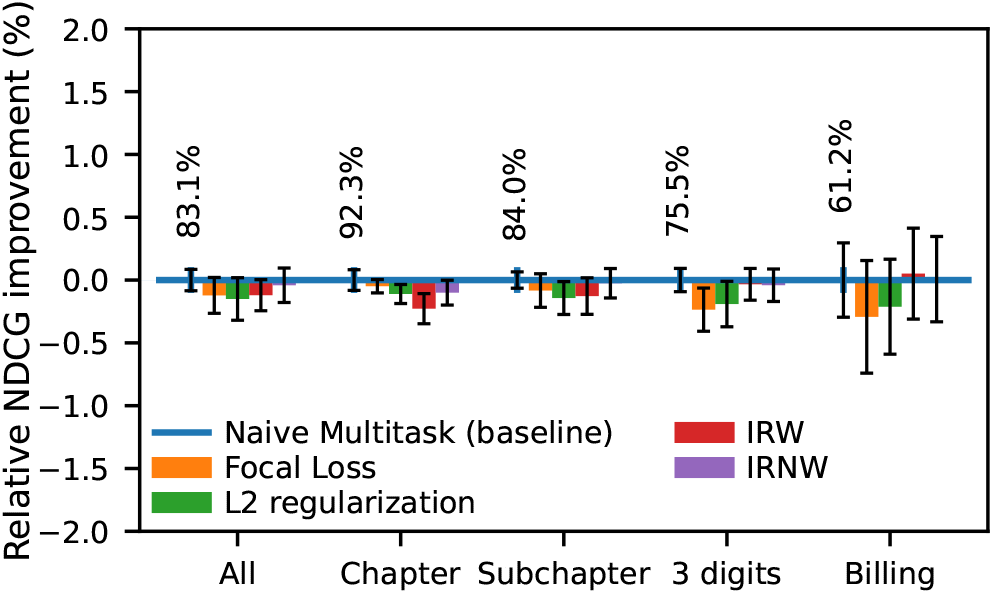
Relative improvement in ranking performance, measured by NDCG, across different levels of the ICD hierarchy using various class imbalance handling strategies compared to the baseline naive multitask model. Absolute NDCG values, averaged over cross-validation runs, for the baseline naive multitask model are provided as reference.

Still, relying on code frequency for ranking purposes raises questions about the models generalization and equity in performance, especially across different wards within the same hospital. To assess the latter, we divided the MIMIC dataset into 3 categories of patients based on patients’ visit details: 8,096 newborn patients, 23,947 surgical patients, and 26,883 medical patients (see Appendix I). Subsequently, we evaluated our model’s performance on each of these patient categories (Table S9). Overall we found that, despite being a minority class, newborn stays exhibited significantly better ranking performance. Moreover, despite the categories having similar frequencies, which may not translate directly into ICD code frequencies, significantly better ranking performance were obtained on the medical stays compared to the surgical ones.

### F. Overall performance summary

#### 1) Predictions

Taken altogether we show in Fig. 6 that our proposed model, with naive multitask learning selected to maximize NDCG, improves the sample averaged Recall@K at all values of K when compared to the strategy similar to the one presented in Hansen et al. [20], the only existing study predicting ICD codes based on medications only. On billing codes average sample recall was increased by 5.22% for K=30 to reach a Recall@30 of 57.05%. On 3-digits codes, the improvement was even greater with 14.77% extra codes retrieved with a Recall@30 of 70.66% (Fig. S7). Beyond ranking, we found that by fine-tuning a model initially optimized for *μ*AUPRC and adjusting the probability threshold at each hierarchy level to maximize *μ*F1 (see Appendix J) we achieved superior *μ*F1 than by direct maximization of *μ*F1 as a target metric (Table S4). Detailed tables presenting ranking and soft classification performance of our models can be found as Table S7 and Table S8.

**Fig. 6:**
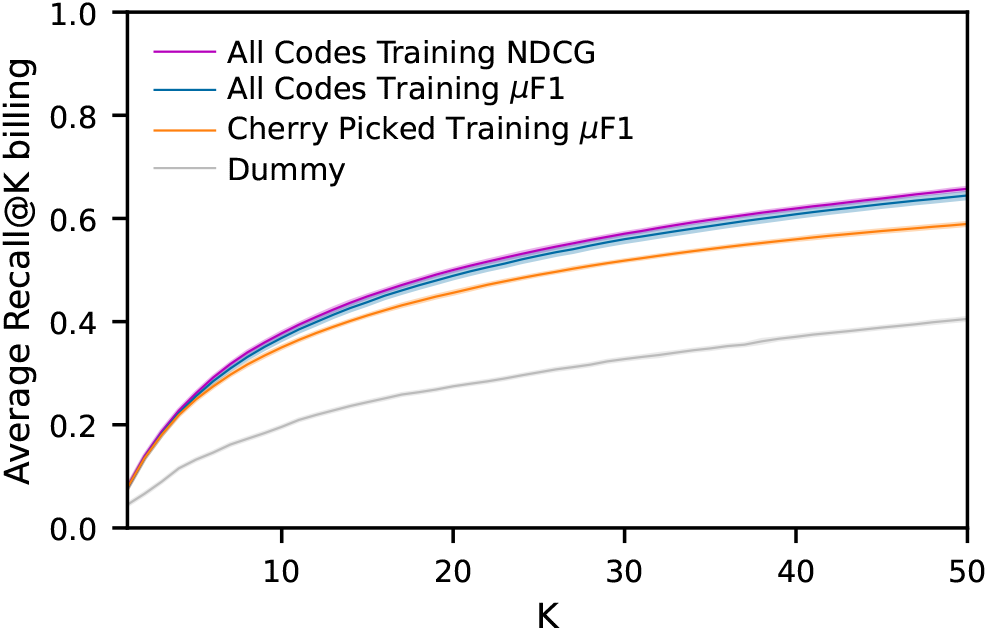
Comparison of ICD code retrieval performance, focused on billing level, between our top-performing model (shown in purple) and a model employing a strategy similar to the state-of-the-art for ICD code prediction using medication data as described in Hansen et al. [20] (represented in orange). Shades around the lines show the standard error on the estimated sample averaged Recall@K computed via three-fold MonteCarlo cross-validation.

#### 2) Runtime

In our extensive experiments and across various cross-validation runs, we consistently observed the emergence of models with similar hyperparameters. Notably, the top-performing models tended to be relatively shallow yet wide networks, featuring 2 to 3 hidden layers, each containing approximately 2-3k units and a dropout rate of approximately 60% (details available in Table S10).

Our model, achieving the best NDCG performance, demonstrated remarkable efficiency in processing the comprehensive MIMIC-III dataset, encompassing 58,976 hospital stays, in 1.92 *±* 0.19 seconds employing an inference batch size of 1024 samples on a T4 NVIDIA GPU.

## IV. Discussion

### A. Choice of an evaluation metric

The first, model-agnostic step towards developing improved CAC systems and encouraging their adoption is the thoughtful selection or definition of a suitable and interpretable performance evaluation metric. To this end, we introduced the RE@R set of evaluation metrics based on existing literature [24, 25] and discussions with health informatics professionals. A previous real-world study [23] showed that a text-based automatic classification model optimized for *μ*F1 improved coding quality but failed to reduce coding time for medical coders. While our proposed evaluation metric fulfills all identified usability and interpretability criteria, further research is needed to validate its usage by studying its correlation with both coding quality and coding time.

### B. Code cherry picking and choice of a target metric

In Section III-A and Section III-B, we demonstrated that filtering ICD codes was detrimental to code retrieval without yielding any positive effects on the model’s performance within the cherry-picked subset. Additionally, using ranking-based metrics, such as NDCG or Precision-Recall curve-derived metrics as proxies to optimize RE@R, proved more effective in selecting superior models compared *μ*F1 . These findings mark a second step toward enhancing ICD CAC systems in a model-agnostic manner. Beyond its adverse impact on code retrieval, code cherry-picking posed challenges for reproducibility, as illustrated by our struggles to replicate filtered code sets across different studies [19, 20, 28]. Although the ranking-based metric NDCG demonstrated the highest correlation with RE@R, our results indicate that AUPRC-based metrics selects better-calibrated models, offering enhanced interpretability for clinical coders. While the observed difference in calibration remained marginal with shallow NN, this distinction might become critical with deep networks, such as text models, which have shown poor calibration without additional correction [37].

### C. Exploiting the ICD hierarchical properties

Our results suggest that minor ranking improvements can be obtained using a naive multitask design (Section III-C). Because the per-level approach requires building one model per hierarchy level, impacting both training and inference time, our result suggest that the naive multitask approach is the most efficient alternative to obtain predictions for different levels of the hierarchy. These results may seem to contradict previous results reporting significant improvement on ICD classification with a multimodal SVM adapted for hierarchical multitask learning [28]. However, the proposed SVM only shares information between explicitly hierarchy-related tasks, while our neural network approach enables multitask learning even using a single hierarchy level through common hidden features [32]. This difference already makes it more akin to explicit HMC methods. Second, the small training set size (3,750 MIMIC-III patients, compared to our 48,976 stays) used in Malakouti and Hauskrecht [28] may have placed their algorithm in a more challenging learning situation where hierarchical multitask learning would be more beneficial. Overall, using our naive multitask neural network approach we obtained much higher classification performances, with 11,64% macro AUROC and 11.72% macro AUPRC differences on comparable code subsets, using only medication information compared to the best reported multimodal SVM in Malakouti and Hauskrecht [28] (see Table S6).

In evaluating the explicit use of the ICD hierarchy (Section III-D), our results indicate that HMC methods did not yield improvements.This contrasts with reports of substantial gains using HMC methods with neural networks [33, 34], in particular for ICD classification using medication data [20]. For the latter, we found that our model’s classification performance was generally superior to results reported by Hansen et al. [20], as discussed in the next subsection. Moreover, their hierarchical strategy, where parent node predictions are set to be the average of children nodes’ predicted probabilities, inherently creates hierarchical violations. This design forces parent node probabilities to be inferior to the most probable child node. We hypothesize that their hierarchical loss formulation promoted a compensatory inflation of leaf node probabilities, resulting in higher recall at the expense of precision and possibly calibration, ultimately contributing to a generally improved *μ*F1 . Regarding other proper HMC methods, Giunchglia and Lukasiewicz [34] reported positive results over several dataset with structured input. However the training set size in their experiments was generally much lower than our setting, on the order of ∼1.5K examples compared to our ∼50k examples. We hypothesize that while HMC methods could show benefits on small datasets, this advantage might diminish with increasing dataset size. Through multitask learning NN are able to learn implicit hierarchical representations [32]. This implicit hierarchical learning could lead to an implicit hierarchy rewiring and ultimately compensate for hierarchical ontological inconsistencies, that would be enforced by an explicit HMC method [36]. The quality of the ICD9 ontology could thus constitute an alternative hypothesis for this lack of improvement using HMC methods.

Regarding class imbalance (Section III-E), our results suggest that our neural network models exploit both input information and code frequency to build predictions without negative impact on minority classes. Furthermore, we observed no added performance gains, from a ranking perspective, when attempting to correct for label imbalance through various approaches with low computational overhead. While class imbalance correction techniques are frequently applied for classification purposes, their exploration within the recommender system domain has been limited. In general, these correction techniques might only alter the position of the decision plane, leaving the embedding space unaffected and resulting in identical ranking performance. The absence of improvement in ranking performance may also stem from the intrinsic multitask nature of recommender systems when utilizing neural networks, which could be particularly beneficial for rare labels.

### D. Runtimes and performance of simple medication based models on MIMIC-III

Our sequential NN model can process medication data with a very high throughput (Section III-F2). Comparatively, Gao et al. [10] reported runtimes for Convolutionnal Neural Network (CNN) and Transformer text models. These models processed 1000 samples in 8.40 and 75.86 seconds, respectively, using a comparable V100 NVIDIA GPU, and focusing on a single level of the hierarchy. In the same experimental conditions, our model exhibited a considerable speed advantage, completing predictions for 1000 samples in 0.077 *±* 0.001 seconds. In addition to being faster by several orders of magnitude, our model output predictions for the entire hierarchy of ICD codes. This runtime difference underscores the significance of utilizing a simple feed-forward NN architecture for conducting our systematic assessment of multitask, hierarchical, and class imbalance correction techniques. Employing more computationally intensive architectures could have rendered this systematic evaluation computationally intractable.

Regarding classification performance using medication data alone, our naive multitask model surpassed the classification results on the cherry-picked code subset reported in Hansen et al. [20] at every level of the hierarchy, except for the 3-digit level (refer to Table S4).

Our results also have to be put in perspective with other approaches using different input data. For instance, Rodrigues-Jr et al. [19] report a 58% Recall@20 using ICD codes history. However this result was obtained on a subset of 855 ICD codes which we were unable to replicate. Additionally, their method is limited to patients who have been previously hospitalized in the database. Our model seemed to be generally surpassed by deep Natural Language Processing (NLP) models that leveraged medical text data [10, 12, 13, 27] on F1 and AUROC based metrics. It’s worth noting that these NLP models were typically evaluated on a broader range of ICD data, encompassing both diagnoses and procedures, making direct comparisons with our work challenging. However, in a noteworthy exception, Gao et al. [10] reported significantly higher *μ*F1 scores using a CNN text model for 3 and 5-digit codes compared to our adaptive threshold model (Table S1 and Table S4). From a computational intensity perspective, though, our feed forward NN aligns more closely with a text-based SVM model employing a bag of words as input [5]. In that study, the SVM achieved a reported *μ*F1 of 29.3% on billing codes. In contrast, our adaptive threshold model exhibits a considerably superior hard classification performance with a 38.04% *μ*F1 on billing codes. This performance difference, with a similarly complex model, underscores the valuable information contained in medication data for ICD coding and encourages further exploration of enhanced model architectures.

We conducted our systematic analysis of plug-in methods to enhance ICD coding using the MIMICIII database, a widely utilized real-world ICU clinical dataset. This choice was motivated by its availability, extensive benchmarks against other algorithms, and the facilitation of reproducibility in our work. Although the reliance on a single dataset might initially seem to restrict the generalization of our results, particularly given its use of ICD-9 labels instead of the more contemporary ICD10, prior studies employing text-based models [17, 23], diagnosis history [19], and even medication data [20] have demonstrated generalization with similar performance across ICD-10[17, 20, 23] variants and other clinical ontologies [19], both on national databases [20] and hospital wide settings [17, 19, 23], when compared to identical architectures trained and evaluated on MIMIC-III. Moreover, we utilized the OMOP-CDM representation of MIMIC-III data to enhance transferability to a real hospital context. Still, it’s important to note that MIMIC-III and other retrospective real-world databases likely contain numerous ICD labeling errors, which may in turn impact our model’s performance evaluation.

## V. Conclusions

In this study, we have explored the construction of more effective CAC systems using generic approaches, with medication data and a simple neural network architecture as our illustrative example. We demonstrated that the practice of ICD code cherry-picking tends to reduce retrieval performance without significant gains in the cherry-picked subset. This not only compromises performance but also poses challenges for reproducibility in research. Our investigation, introducing a novel metric RE@R tailored for the ICD coding task, revealed that optimizing for metrics like NDCG and AUPRC leads to superior ranking performance compared to the commonly used F1-based metrics. We found that multitask learning, by training NNs simultaneously on different ICD hierarchy levels, provides minor benefits for ranking as well as runtime gains. Surprisingly, our sequential NN models did not draw benefits from HMC methods or class imbalance correction techniques.

Our generic experiments offer valuable insights for researchers and healthcare practitioners interested in developing or evaluating CAC systems. Despite employing a straightforward sequential NN model, we confirmed that medical prescriptions represent a rich data source for CAC systems, providing competitive ICD codes retrieval capabilities for a fraction of the computational load compared to text-based models. Our conclusions however, bear the limitations inherent to the dataset. First and foremost, our conclusions regarding the use of the ICD hierarchy are based on ICD9, while ICD10 is the current standard, featuring extensive reorganization and an increased number of labels. Furthermore, MIMIC-III contains only ICU stays, which might not fully represent the diversity of medical practices, and potential ward balance issues could arise on a hospital level despite our reassuring initial experiments. Our study employed a straightforward sequential neural network model to demonstrate the potential of medication data for CAC systems. Future work could explore more sophisticated architectures that leverage additional information, such as dosage, route of administration, and temporal patterns in medication intake, to further enhance the prediction quality.

## Data Availability

All data produced in the present work are contained in the manuscript. Software will be made available at https://github.com/qmarcou upon acceptance of the manuscript.

## Authors’ contributions

**QM:** Conceptualization, Methodology, Software, Formal analysis, Investigation, Visualization, Writing Original Draft, Writing - Review & Editing. **LB and NN:** Supervision, Writing - Review & Editing.

## Declaration of interests

The authors declare that they have no known competing financial interests or personal relationships that could have appeared to influence the work reported in this paper.

Acknowledgments

V. Pradel, F. Antonini and C. Fraboulet for initial discussions. M. Bertrand for the support of the LIS computing infrastructure.

## Code availability

Computer code will be released under a FOSS license on QM’s Github profile: https://github.com/qmarcou

## Acronyms

AUPRC: Area Under the Precision Recall Curve.
AUROC: Area Under the Receiver Operating Characteristic Curve.
BCE: Binary Cross-Entropy.
CAC: Computer Assisted Coding.
CNN: Convolutionnal Neural Network.
ECE: Expected Calibration Error.
HMC: Hierarchical Multilabel Classification.
ICD: International Classification of Diseases.
IRNW: Imbalance Ratio Normalized Weighting.
IRW: Imbalance Ratio Weighting.
MCE: Maximum Calibration Error.
MTL: Multitask Learning.
NDCG: Normalized Discounted Cumulative Gain.
NLP: Natural Language Processing.
NN: Neural Network.
RNN: Recurrent Neural Network.
SVM: Support Vector Machine.

## Appendix

### A. Notations

- *N* the number of training examples
- *M* the number of labels, or nodes, in the hierarchy
- y the vector of true labels
- *y*_*i*_ the true value of label *i*
- *ŷ* the vector of predictions
- *ŷ*^+^ the vector of predictions for positive labels
- *ŷ*^−^ the vector of predictions for negative labels
- 𝒟_*i*_ the set of descendants of node *i*
- 𝒜_*i*_ the set of ancestors of node *i*
- π (*i*) the parent of node *i*

### B. Usual performance metrics and averaging strategies

We report the following standard metrics based on confusion matrices: namely Precision, Recall, and some variants of their harmonic mean the F1 score, as well as areas under the Precision-Recall curve (AUPRC) and area under the Receiver Operating Characteristic curve (AUROC).

Micro-averaged metrics denote metrics computed discarding class and sample belonging information, meaning each prediction-value pair is treated equally. Sample-averaged metrics denote metrics computed first on a per example basis and then averaged over examples.Macro metrics on the other hand denote metrics computed first on a per label basis and then averaged over labels. Macro metrics will penalize more severely models with good performance on some frequent labels but poor performance on less frequent or more difficult labels.

All the performance metrics that we report throughout the paper are ill-defined when no positive example is present in the evaluated dataset. This problem arises upon performing sample or macro averaging. For sample averaging, some hospital stays have no associated ICD codes. For macro averaging, several rare codes are too rare to be seen in a test set subsample, the number of such codes per test set is dependent on the chosen test set size. In order to minimize the bias that could be introduced by assigning a default value (such as 0 or 1) to a metric when no positive example is in the dataset, we ignored stays or codes without positive example during sample or macro averaging. Note however that for macro averaging this strategy tends to leave out very rare labels if the test set size is too small, and thus results in an optimistic bias.

### C. Rank-based metrics

#### 1) Normalized Discounted Cumulative Gain (NDCG)

NDCG is the normalized version of Discounted Cumulative Gain (DCG) and is a widely used ranking metric. NDCG is defined as:

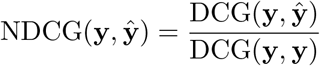

where DCG(y, y) is the DCG value for an ideal ranking.

DCG exists in several versions with different gain and rank discount functions. We chose the most common DCG fomulation, with 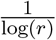 discount. [S1]. Within our binary relevance framework, we thus formulate DCG as:

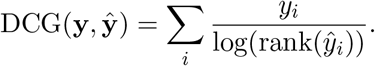

We did not restrict NDCG computation to a range of top K predictions (NDCG@K) in order to capture differences in models performances, even for incorrectly ranked positive labels. Unless stated otherwise we computed the NDCG using the set of codes appearing at least once in MIMIC-III, or one of its hierarchical subsets. Note that due intrisic properties of NDCG, computing NDCG using the complete ICD-9, thus including codes that did not appear in the MIMIC-III database, would tend to produce values closer to 1 [S2].

#### 2) Coverage

Coverage measures how far on the ranked predictions list ones has to go to find back every positive label:

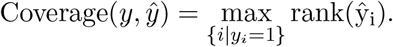

### D. Calibration Error

The expected calibration error measures the adequacy between the confidence of the model in its prediction and the accuracy of predictions for that level of confidence [S3]. We computed the ECE and MCE using an adaptive binning strategy with 500 bins.

### E. Loss function

#### 1) Cross entropy loss

We treat the multilabel classification by treating each label as a separate binary classification task in a final dense layer, while using a common se t of layers performing feature extraction. This can be done using the Binary cross-entropy or logistic loss function:

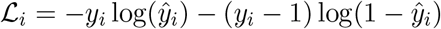

#### 2) Focal loss

The Binary Cross-Entropy (BCE) can be altered to put more emphasis on “difficult” classification samples, resulting in the focal loss [S4]:

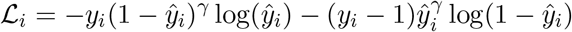

where *γ* is a tunable hyperparameter.

#### 3) Re-weighted multitask

In order to give equal weight to each hierarchy level in the loss function, we can reweight each label by the inverse proportion of labels of that level in the hierarchy:

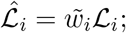

where 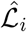 is our modified loss function and 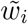 a weighting factor. We defined *w*_*i*_, the inverse proportion of labels in the hierarchy in the same level as label *i* as:

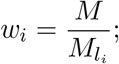

and its normalized version 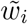, such that the average level weight is 1:

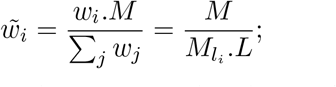

where 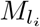 is the number of nodes in the hierarchy level *l*_*i*_ and *L* the number of levels in the hierarchy.

For our experiment we considered 4 different levels: Chapters, Subchapters, 3 digits and a final level grouping both 4 and 5 digits levels, to reflect the fact that Billing codes of interest can be either 3, 4 or 5 digits codes.

### F. Hierarchical multilabel classification methods

Hierarchical classification is by nature multilabel (since a label from a leaf entails the parent label). Here by HMC we denote the fact that each example can be labeled by several labels from the same level in the hierarchy. HMC can be tackled with 3 different approaches: flat classifier (that we refer to as naive multitask model in main text), local classifiers and global classifier [S5]. Flat classification is the simplest approach and simply ignores the hierarchy information thus training an ensemble of binary classifiers (possibly including non leaf nodes) to perform a simple multilabel classification.

The local classifier approach consists in training different classifiers for different levels in the hierarchy. The resulting number of classifier can vary according to the chosen approach (Local classifier per node, local classifier per parent, local classifier per level). In the local classifier approach, one uses the prediction from the parent classifier to interpret the result from the child classifier. The intuitive advantage of building several classifiers being that it could, in theory, allow the algorithm to pick different features scales for decision making. On the contrary the obvious drawback is in the need to build many classifiers, meaning longer training time, harder evaluation, and longer inference time.

Finally the global classifier approach aims at building a single classifier for all hierarchy levels, and exploit the hierarchy information via a dedicated algorithm structure, regularization [S6] enforcing similarity between parent and child nodes (and thus implicitly among siblings) or by introducing a penalty based on tree distance inside the cost function [S7]. Some recent papers enforce logical constraints on hierarchical coherence of predictions at learning time inside the cost function [S8, S9] Finally in some recent work [S10] tried to bridge the gap and propose a hybrid approach, between the global and the local one, based on neural networks.

Hierarchy inconsistencies in predictions of global classifiers (predicting a lower probability for a parent node than for one of its children) can be limited by a regularization [S10] or cost [S8, S9] approach at learning time, and can be further enforced through imposing logical constraints at inference time [S8].

In our work, we focused on global approaches, that are model architecture agnostic, since most existing work on ICD assignment use a simple multilabel framework.

#### 1) Hierarchical logical constraints

Upon training a flat or global classifier some output predictions may violate the hierarchy. Such violations can be reduced at inference time by introducing simple logical constraints such as roll-up:

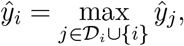

or roll-down:

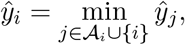

of the hierarchy predictions. As discussed in [S8], the choice of the correct strategy will depend on the actual problem at hand and the topological structure of label ensembles in the feature space. However, by imposing such constraints at inference time only, the hierarchy may only be used for hyper-parameter tuning, but not for the actual training of the model. To alleviate this limitation some recent works introduce such logical constraints into the loss function. In [S8] the authors introduce the MCLoss (Maximum Constraint Loss),a variant of the binary cross entropy:

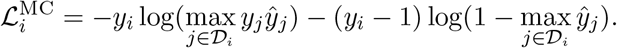

This choice of logical constraint, together with a roll-up at inference time, is motivated by a discussion on the optimal hierarchical classification strategy according to the topology of labels in the feature space. Using the MCLoss and roll-up, the classifier should self adapt to the actual topology of labels in the feature space and always behave like the best classifying strategy [S8].

In [S9] introduce the TreeMin loss:

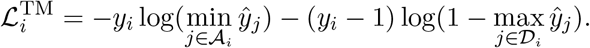

This loss can be intuitively understood as always penalizing according to the worst relevant prediction in the hierarchy. One advantage of these logical constraint approaches is that they do not require to introduce new hyperparameters to be tuned.

#### 2) Hierarchical regularization

Another way to use the hierarchical relationships is to use the notion that labels close in the hierarchy should also be close in the feature space.

A simple way to leverage this idea is to introduce a regularization term in the loss function enforcing similarity of the parameters between children and parent nodes [S6]:

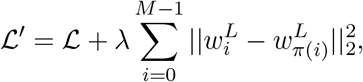

where *λ* is a tunable hyperparameter.

### G. Class imbalance

In general methods to handle class imbalance can be decomposed in three families: cost-sensitive or classifier adaptation, ensemble learning, and resampling, with the latter one being predominant due to its applicability to any classifier type [S11]. Multilabel classification already adds complexity to the imbalance problem by the fact that by resampling one might also change the frequency of some associated labels, and specific methods have been designed to address this issue. [S11, S12].

HMC adds a related problem: each time a leaf is sampled the corresponding parent will be sampled too, making it difficult to control balance for each node in the hierarchy tree. Only a small number of studies try to address the imbalance problem for hierarchical classification. The proposed approaches, however, rely on local classifiers [S13] or transformation to label sets [S14] to perform this resampling. As explained earlier we chose to focus on global classifier approaches using binary relevance such that our results are most easily transferable to existing work on ICD classification. Because some of the models used in ICD codings, such as Transformers, are already computationally demanding we decided to focus on exploring some plug in cost sensitive methods, and leave out ensemble methods.

One of the simplest ways to address imbalance using a cost sensitive approach without an expert pre-specified cost matrix is by increasing the weight of mistakes on positive samples according to each label imbalance ratio (IR):

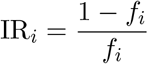

Using the BCE loss function, we can define the IRW loss:

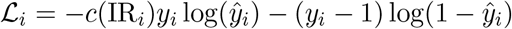

where *c*(IR_*i*_) can be any increasing function of IR such that *c*(IR_*i*_) ≤ IR_*i*_. By strongly penalizing mistakes on positive examples of some labels, we increase their importance during training. This strategy aims at pushing decision boundaries away from the positive instances, and in theory leads to an improved generalization on these labels. The extreme case *c*(IR_*i*_) = IR_*i*_ can be seen as an extreme upsampling like procedure where the total weight of positive examples is equal to the total weight of negative examples. However, because of the limited amount of positive examples for some labels, their importance might be blown up and result in poor generalization and ultimately lead to degradation of performances on negative examples.

In this study, for the IRW method, we defined *c* to be a simple ceiling function:

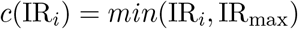

where IR_max_ is a tunable hyperparameter

One caveat of such a re-weighting in the multilabel context is that labels end up with different weights in the total loss according to the label frequency. If we consider an adversarial classifier always giving the worst prediction up to *ϵ*, the total loss for a given label *i* is:

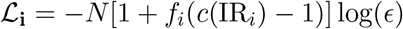

To alleviate this we simply rescale each label contribution:

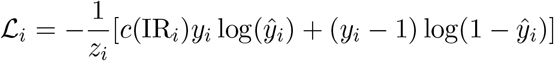

by the normalizing factor *z*_*i*_ = 1 + *f*_*i*_(*c*(IR_*i*_) − 1). In the remainder of the text, we refer to this normalized re-weighting as IRNW.

### H. Entropy reduction

We calculate the entropy reduction, also known as information gain, by analyzing the predictions made by our model on the test set. Initially, we compute the mutual information between the model’s predictions and the ground truth of the test set using the ‘mutual_info_classif’ function from the ‘sklearn’ library, considering continuous predictions for each cross-validation run. Due to the limited size of the test sets, we encounter some codes for which no positive examples were observed in some or all test sets; in such cases, the mutual information is set to NaN.

To obtain the entropy reduction, we normalize the mutual information by the entropy of the code, which is estimated using the entire dataset. For 36 codes, the resulting entropy reduction values exceeded 1.0; we capped these values to 1.0 to ensure consistency.

### I. Stay types categorization

In Section III-E, we assess our model’s performance across distinct stay categories: newborn, surgical, and medical.

To categorize each stay, we utilized the *visit_detail* table within MIMIC-OMOP and classified stays based on the *visit_detail_concept_id* field. Stays containing at least one entry corresponding to the newborn category (concept 4237225) were assigned to that group. Similarly, stays lacking newborn category assignment but having at least one surgical detail entry (concept 4149152) were categorized as surgical stays. For the remaining stays, they were considered medical stays provided they had the corresponding entry (concept 45763735).

Notably, 50 stays from the entire database could not be assigned to any of these categories and were consequently excluded from this analysis, provided they appeared in a test set.

### J. Adaptive threshold model

Since the cross-entropy loss is not a direct optimizer for *μ*F1, models selected based solely on *μ*F1 via hyperparameter search and early stopping may not deliver the optimal results in terms of hard classification. To enhance the hard classification results, as measured by *μ*F1, we adopted models initially optimized for *μ*AUPRC. We then fine-tuned these models by adjusting the probability threshold for each hierarchy level to classify examples as positive or negative. These thresholds were carefully chosen to maximize *μ*F1 on the 5,000 stays from the development dataset. Results on the cherry-picked code set are given in Table S4.

### K. Runtime evaluation

To evaluate runtime we used the best model optimizing NDCG on the first cross validation dataset. We conducted evaluations across multiple batch sizes, ranging from 32 to 16,296, aiming to enhance inference efficiency on the complete dataset comprising 58,976 stays. In order to eliminate the compilation time overhead of the model, we initially conducted inference on a single batch. Subsequently, we performed inference on both the complete dataset and a random subset of 1,000 stays, timing the process over five repetitions. Our reported results represent the average runtime, based on these five runs, along with the corresponding standard error. For achieving the best performance on the complete dataset, we selected a batch size of 4,096. This experiment was executed on Google Colab, utilizing a T4 GPU

### L. Supplementary tables

**TABLE S1:**
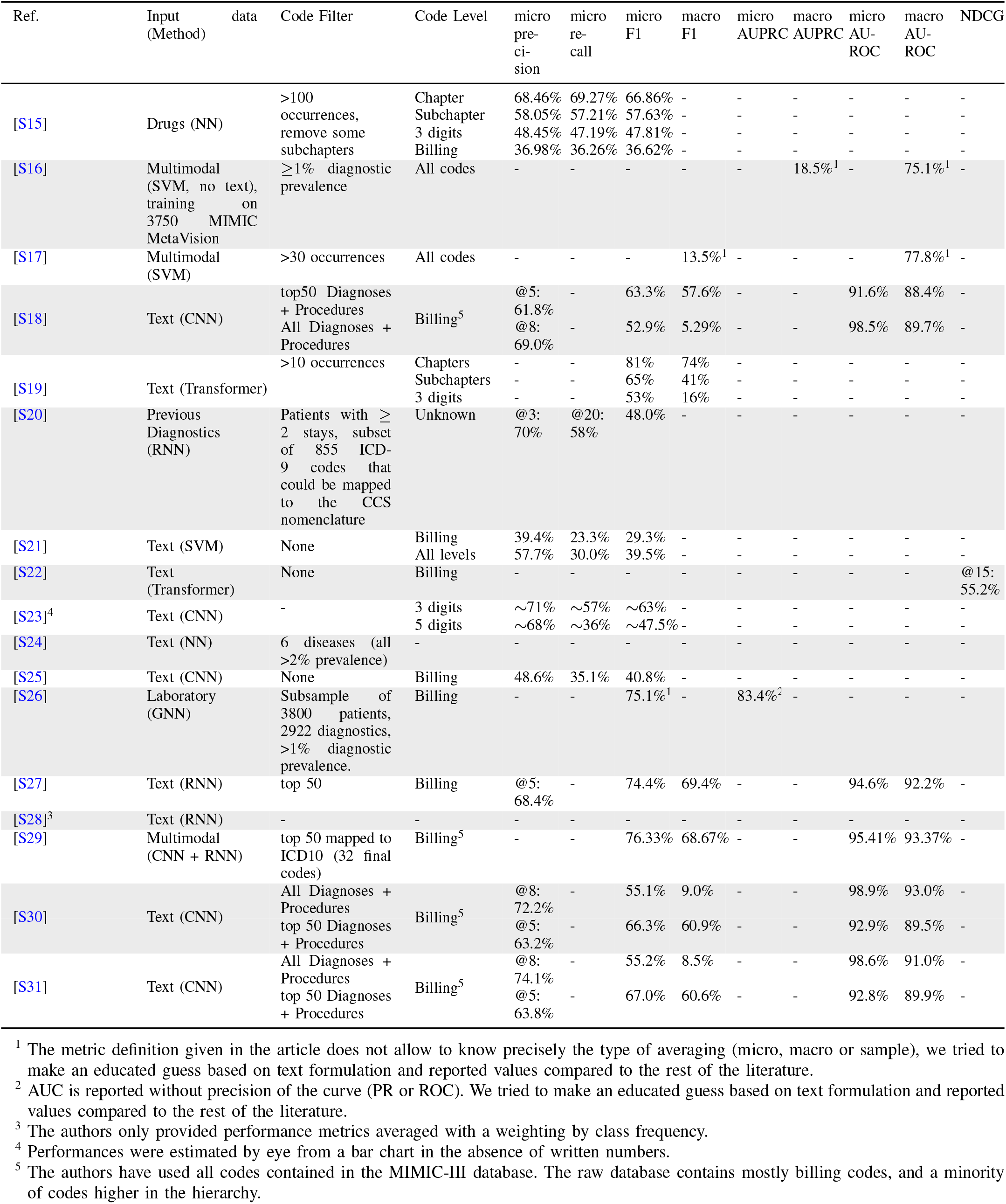
Summary of existing work on ICD9 prediction on MIMICIII.

| Ref. | Input data | Code Filter | Code Level | micro pre-cision | micro re-call | micro F1 | macro F1 | micro AUPRC | macro AUPRC | micro AU-ROC | macro AU-ROC | NDCG |
| --- | --- | --- | --- | --- | --- | --- | --- | --- | --- | --- | --- | --- |
| [S15] | Drugs (NN) | >100 occurrences, remove some subchapters | Chapter<br>Subchapter<br>3 digits<br>Billing | 68.46%<br>58.05%<br>48.45%<br>36.98% | 69.27%<br>57.21%<br>47.19%<br>36.26% | 66.86%<br>57.63%<br>47.81%<br>36.62% | -<br>-<br>-<br>- | -<br>-<br>-<br>- | -<br>-<br>-<br>- | -<br>-<br>-<br>- | -<br>-<br>-<br>- | -<br>-<br>-<br>- |
| [S16] | Multimodal (SVM, no text), training on 3750 MIMIC MetaVision | $\geq 1\%$ diagnostic prevalence | All codes | - | - | - | - | - | 18.5%<sup>1</sup> | - | 75.1%<sup>1</sup> | - |
| [S17] | Multimodal (SVM) | >30 occurrences | All codes | - | - | - | 13.5%<sup>1</sup> | - | - | - | 77.8%<sup>1</sup> | - |
| [S18] | Text (CNN) | top50 Diagnoses + Procedures<br>All Diagnoses + Procedures | Billing<sup>5</sup> | @5: 61.8%<br>@8: 69.0% | - | 63.3%<br>52.9% | 57.6%<br>5.29% | -<br>- | -<br>- | 91.6%<br>98.5% | 88.4%<br>89.7% | -<br>- |
| [S19] | Text (Transformer) | >10 occurrences | Chapters<br>Subchapters<br>3 digits | -<br>-<br>- | -<br>-<br>- | 81%<br>65%<br>53% | 74%<br>41%<br>16% | -<br>-<br>- | -<br>-<br>- | -<br>-<br>- | -<br>-<br>- | -<br>-<br>- |
| [S20] | Previous Diagnostics (RNN) | Patients with $\geq 2$ stays, subset of 855 ICD-9 codes that could be mapped to the CCS nomenclature | Unknown | @3: 70% | @20: 58% | 48.0% | - | - | - | - | - | - |
| [S21] | Text (SVM) | None | Billing<br>All levels | 39.4%<br>57.7% | 23.3%<br>30.0% | 29.3%<br>39.5% | -<br>- | -<br>- | -<br>- | -<br>- | -<br>- | -<br>- |
| [S22] | Text (Transformer) | None | Billing | - | - | - | - | - | - | - | - | @15: 55.2% |
| [S23]<sup>4</sup> | Text (CNN) | - | 3 digits<br>5 digits | $\sim 71\%$<br>$\sim 68\%$ | $\sim 57\%$<br>$\sim 36\%$ | $\sim 63\%$<br>$\sim 47.5\%$ | -<br>- | -<br>- | -<br>- | -<br>- | -<br>- | -<br>- |
| [S24] | Text (NN) | 6 diseases (all >2% prevalence) | - | - | - | - | - | - | - | - | - | - |
| [S25] | Text (CNN) | None | Billing | 48.6% | 35.1% | 40.8% | - | - | - | - | - | - |
| [S26] | Laboratory (GNN) | Subsample of 3800 patients, 2922 diagnostics, >1% diagnostic prevalence. | Billing | - | - | 75.1%<sup>1</sup> | - | 83.4%<sup>2</sup> | - | - | - | - |
| [S27] | Text (RNN) | top 50 | Billing | @5: 68.4% | - | 74.4% | 69.4% | - | - | 94.6% | 92.2% | - |
| [S28]<sup>3</sup> | Text (RNN) | - | - | - | - | - | - | - | - | - | - | - |
| [S29] | Multimodal (CNN + RNN) | top 50 mapped to ICD10 (32 final codes) | Billing<sup>5</sup> | - | - | 76.33% | 68.67% | - | - | 95.41% | 93.37% | - |
| [S30] | Text (CNN) | All Diagnoses + Procedures<br>top 50 Diagnoses + Procedures | Billing<sup>5</sup> | @8: 72.2%<br>@5: 63.2% | -<br>- | 55.1%<br>66.3% | 9.0%<br>60.9% | -<br>- | -<br>- | 98.9%<br>92.9% | 93.0%<br>89.5% | -<br>- |
| [S31] | Text (CNN) | All Diagnoses + Procedures<br>top 50 Diagnoses + Procedures | Billing<sup>5</sup> | @8: 74.1%<br>@5: 63.8% | -<br>- | 55.2%<br>67.0% | 8.5%<br>60.6% | -<br>- | -<br>- | 98.6%<br>92.8% | 91.0%<br>89.9% | -<br>- |

<sup>1</sup> The metric definition given in the article does not allow to know precisely the type of averaging (micro, macro or sample), we tried to make an educated guess based on text formulation and reported values compared to the rest of the literature.
<sup>2</sup> AUC is reported without precision of the curve (PR or ROC). We tried to make an educated guess based on text formulation and reported values compared to the rest of the literature.
<sup>3</sup> The authors only provided performance metrics averaged with a weighting by class frequency.
<sup>4</sup> Performances were estimated by eye from a bar chart in the absence of written numbers.
<sup>5</sup> The authors have used all codes contained in the MIMIC-III database. The raw database contains mostly billing codes, and a minority of codes higher in the hierarchy.

**TABLE S2:** Number of codes per stay for different ICD levels on the cherry picked code subset.

|  | Min | Max | Median | Mean | SD |
| --- | --- | --- | --- | --- | --- |
| Chapters | 0 | 14 | 4 | 4.52 | 2.61 |
| Subchapters | 0 | 24 | 6 | 6.12 | 3.91 |
| 3 digits | 0 | 32 | 7 | 7.83 | 4.98 |
| 4 digits | 0 | 34 | 7 | 7.43 | 5.17 |
| 5 digits | 0 | 22 | 3 | 3.18 | 2.72 |
| Billing | 0 | 34 | 7 | 7.43 | 5.23 |
| All levels | 0 | 112 | 27 | 29.07 | 18.57 |

**TABLE S3:** Models’ Hyperparameters space.

| Hyperparameter | Value | Allowed values |
| --- | --- | --- |
| Global parameters |  |  |
| Test set size | 5000 |  |
| Dev set size | 5000 |  |
| Network parameters |  |  |
| Batch size | 64 |  |
| Number of layers |  | [1, 8] |
| Layers size |  | [256, 6144] |
| Hidden layers activation | ReLU |  |
| Dropout rate |  | [0, 80%] |
| BatchNorm |  | {True, False} |
| Optimization parameters |  |  |
| ADAM: $\beta_1$ | 0.9 | |
| ADAM: $\beta_2$ | 0.999 | |
| ADAM: $\epsilon$ | $10^{-7}$ | |
| ADAM: Learning rate | $2 \times 10^{-4}$ | |
| Earlystopping: patience | 3 |  |
| Earlystopping: min $\Delta$ | 0.0 | |
| Earlystopping: restore best weights | True |  |
| Earlystopping: validation split | 5% |  |
| Hyperparameter search parameters |  |  |
| Hyperband: Iterations | 2 |  |
| Hyperband: Execution per trial | 3 |  |
| Hyperband: Factor | 3 |  |
| Hyperband: Max epochs | 150 |  |
| Method specific parameters |  |  |
| Imbal: Class weight cap |  | [1, 200] |
| Focal: $\gamma$ | | [1, 4] |
| HierL2: $\lambda$ | | $[10^{-15}, 5^{-6}]$ |
| OutL2: $\lambda$ | | $[10^{-8}, 10^{-2}]$ |

**TABLE S4:**
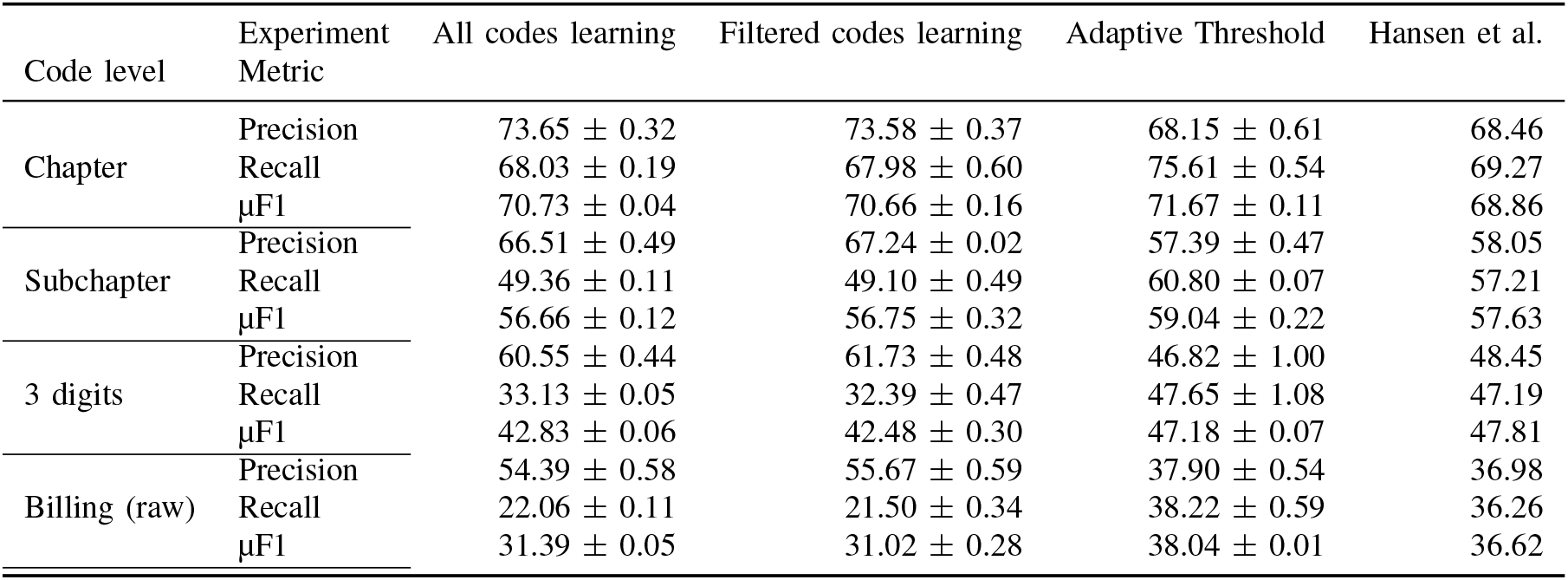
Models classification performance comparison on the cherry picked codes. All codes learning and Filtered code learning designate models that were selected to maximize *μ*F1 and were trained using the complete code set or using only cherry picked codes. The Adaptive threshold model is a model initially trained and selected to maximize *μ*AUPRC, for which a different probability threshold for each level of the hierarchy in order to maximize *μ*F1 (see Appendix J). For comparison we report the performance from Hansen et al. [S15] which we tried to emulate. For our experiment we report the standard error on the mean over the 3 cross-validation runs. Despite our efforts we could not exactly replicate the filters from Hansen et al. [S15], and our cherry picked code set contains ∼100 more codes at the 3-digits and billing levels than that reported in their article.

**TABLE S5:** Calibration metrics for models selected to optimize respectively the AUPRC and NDCG. The table reports mean and standard error on the mean evaluated using 3 cross-validation runs.

| Calibration Metric<br>Optimized Metric | Brier score | ECE | MCE |
| --- | --- | --- | --- |
| AUPRC | 0.282 ± 0.00% | 0.039 ± 0.00% | 1.171 ± 0.23% |
| NDCG | 0.284 ± 0.00% | 0.055 ± 0.00% | 2.297 ± 0.33% |

**TABLE S6:** Models classification performance comparison between our naive multitask neural network and results from a multimodal multitask SVM reported in [S16]. Our neural network model was trained and selected to maximize the macro Area Under the Receiver Operating Characteristic Curve (AUROC) on the complete code set using split sizes given in Section II-B1. The reported macro AUROC and AUPRC were evaluated on a subset of 592 codes with frequency ≥1% on the complete MIMIC-III dataset. In turn, the SVM results given in [S16] were obtained after training on a 3750 patients subset of the Metavision MIMIC subset, with the same code frequency threshold however resulting in 692 codes. For our experiment we report the standard error on the mean over the 3 cross-validation runs.

| Metric | Naive multitask (592 codes) | Malakouti and Hauskrecht (696 codes) |
| --- | --- | --- |
| macro AUPRC | 30.22 ± 0.15 | 18.5 |
| macro AUROC | 86.74 ± 0.05 | 75.1 |

**TABLE S7:** Average ranking or retrieval performance, along with its standard error, estimated on diverse code subsets using three-fold Monte-Carlo cross-validation. These metrics were computed using our top-performing models selected to optimize NDCG.

| Filter | Metric Hierarchy level | NDCG | NDCG@15 | RE@25 | RE@50 | RE@75 | RE@100 |
| --- | --- | --- | --- | --- | --- | --- | --- |
| None | Chapter | 92.33 ± 0.07 | 91.71 ± 0.11 | 0.404 ± 0.00 | 0.987 ± 0.01 | 2.134 ± 0.01 | 4.094 ± 0.01 |
| | Subchapter | 83.98 ± 0.05 | 73.81 ± 0.07 | 1.719 ± 0.04 | 4.196 ± 0.09 | 9.977 ± 0.12 | 26.47 ± 0.22 |
| | 3 digits | 75.52 ± 0.07 | 59.61 ± 0.05 | 5.516 ± 0.16 | 15.07 ± 0.25 | 39.99 ± 0.30 | 148.4 ± 1.54 |
| | Billing | 61.23 ± 0.18 | 43.96 ± 0.11 | 19.35 ± 0.26 | 52.29 ± 1.09 | 155.4 ± 2.85 | 813.3 ± 13.0 |
| | All | 83.09 ± 0.07 | 74.68 ± 0.11 | 8.185 ± 0.28 | 37.30 ± 0.53 | 146.4 ± 1.68 | 1696. ± 24.2 |
| Top50 | 3 digits | 78.88 ± 0.14 | 71.27 ± 0.11 | 1.716 ± 0.00 | 3.602 ± 0.01 | 7.218 ± 0.04 | 14.02 ± 0.07 |
| | Billing | 63.95 ± 0.42 | 58.20 ± 0.44 | 2.279 ± 0.06 | 4.423 ± 0.09 | 8.454 ± 0.10 | 15.38 ± 0.12 |
| | All | 90.60 ± 0.06 | 80.32 ± 0.04 | 1.311 ± 0.02 | 3.536 ± 0.04 | 7.819 ± 0.03 | 17.22 ± 0.05 |

**TABLE S8:** Average (soft) classification performance, along with its standard error, estimated on diverse code subsets using three-fold Monte-Carlo cross-validation. These metrics were computed using our top-performing models selected to optimize each of the corresponding metric.

| Filter | Metric Hierarchy level | μAUROC | macro AUROC | μAUPRC | macro AUPRC |
| --- | --- | --- | --- | --- | --- |
| None | Chapter | 89.29 ± 0.06 | 84.67 ± 0.07 | 80.48 ± 0.10 | 68.37 ± 0.67 |
| | Subchapter | 94.87 ± 0.04 | 85.57 ± 0.16 | 62.07 ± 0.14 | 28.90 ± 0.41 |
| | 3 digits | 96.95 ± 0.01 | 85.21 ± 0.24 | 45.22 ± 0.15 | 13.46 ± 0.04 |
| | Billing | 97.59 ± 0.01 | 84.48 ± 0.20 | 29.84 ± 0.01 | 7.303 ± 0.07 |
| | All | 98.30 ± 0.00 | 84.61 ± 0.19 | 49.92 ± 0.12 | 9.493 ± 0.12 |
| Top50 | 3 digits | 89.20 ± 0.08 | 85.66 ± 0.08 | 59.77 ± 0.25 | 49.26 ± 0.17 |
| | Billing | 88.99 ± 0.09 | 85.24 ± 0.12 | 49.25 ± 0.27 | 37.29 ± 0.23 |
| | All | 87.03 ± 0.04 | 84.82 ± 0.05 | 75.28 ± 0.04 | 68.76 ± 0.15 |

**TABLE S9:**
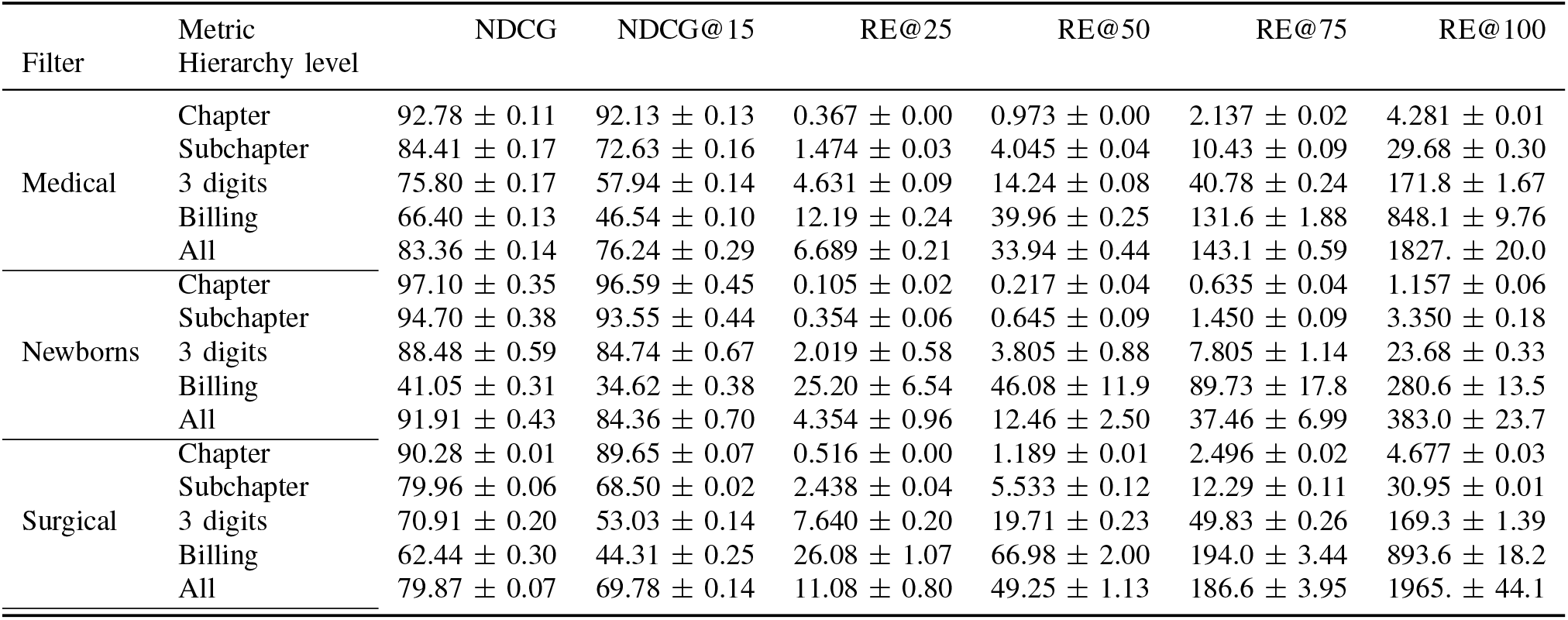
Ranking or retrieval performance across various stay categories. The reported values are averages taken from the test sets of the three cross-validation runs, utilizing models selected to optimize NDCG.

**TABLE S10:** Best models hyper-parameters on the three cross validation runs aiming to optimize NDCG.

| Cross-validation ID | Batch Normalization | Number of Layers | Number of Units | Dropout Rate |
| --- | --- | --- | --- | --- |
| 0 | False | 3 | 2048 | 0.66 |
| 1 | False | 3 | 4608 | 0.63 |
| 2 | False | 3 | 2560 | 0.64 |

### M. Supplementary figures

**Fig. S1:**
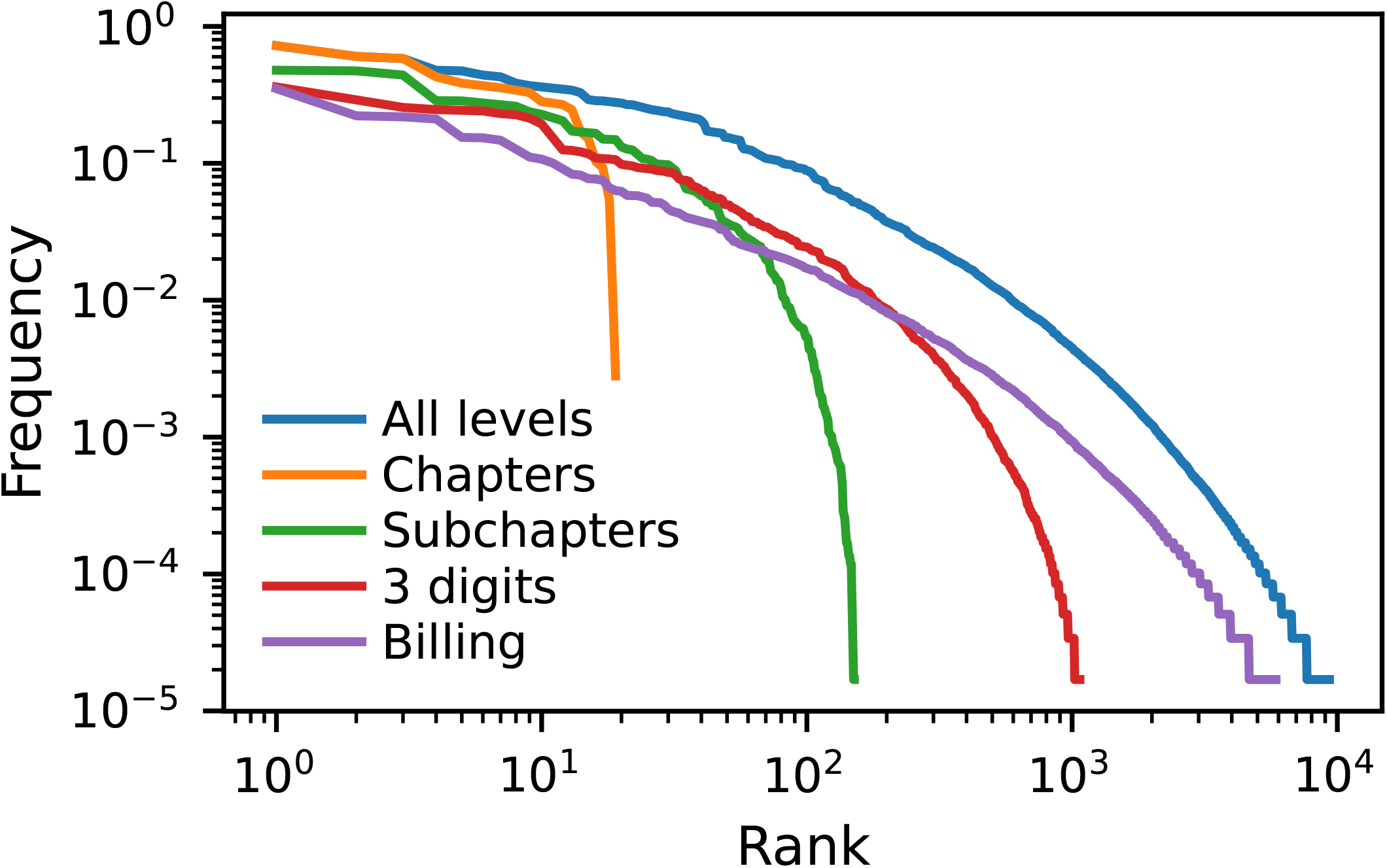
Rank versus frequency plot (Zipf plot) for the different code levels in MIMIC-III.

**Fig. S2:**
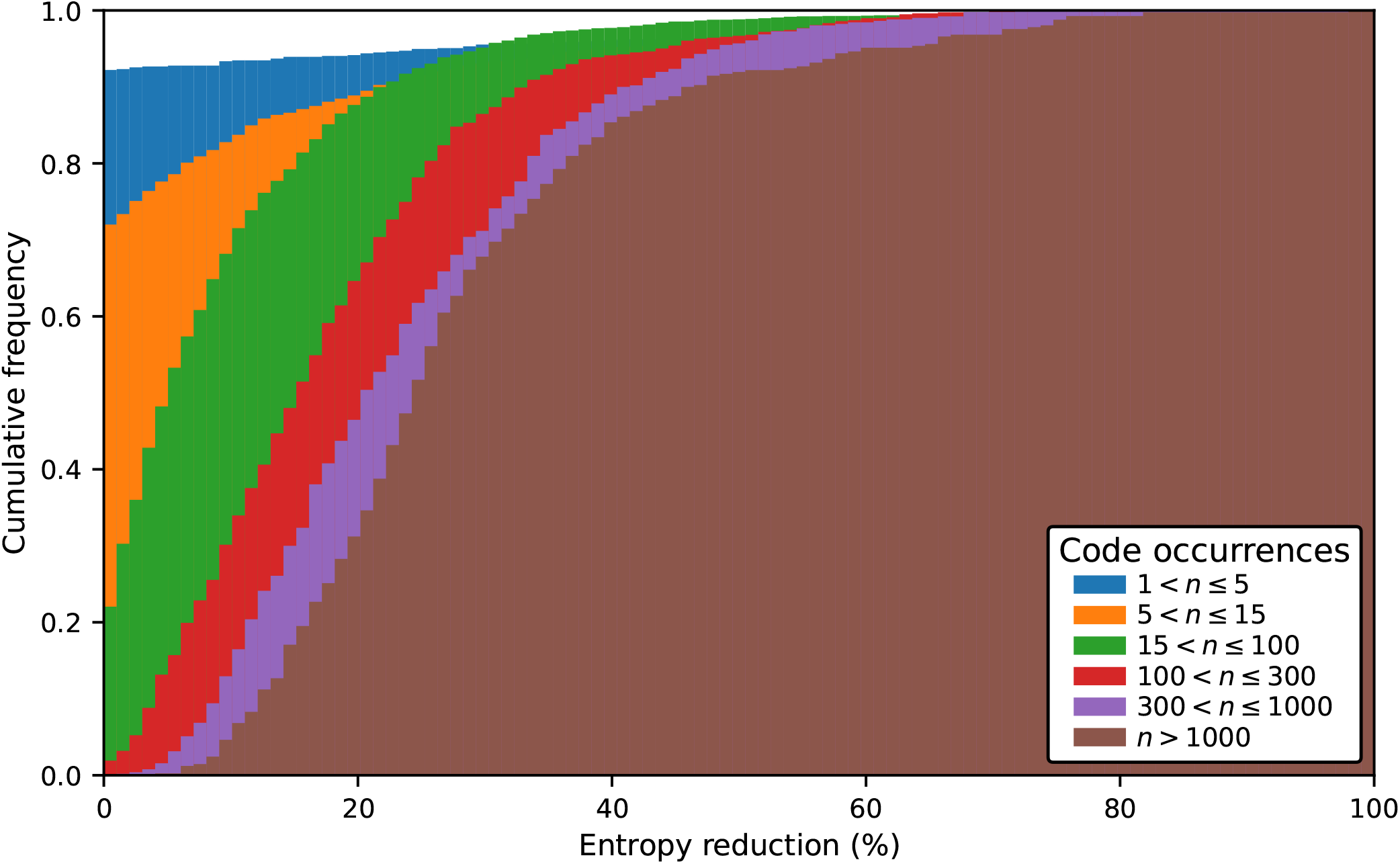
Cumulative distribution of the entropy reduction, or information gain, for different subsets of low frequency codes at any level of the ICD9 hierarchy. Each subset contains respectively, by increasing number of code occurrences, 1350, 1598, 2061, 783, 510 and 410 codes. The results shown were obtained using predictions from a model with hyperparameters selected to achieve best *μ*F1.

**Fig. S3:**
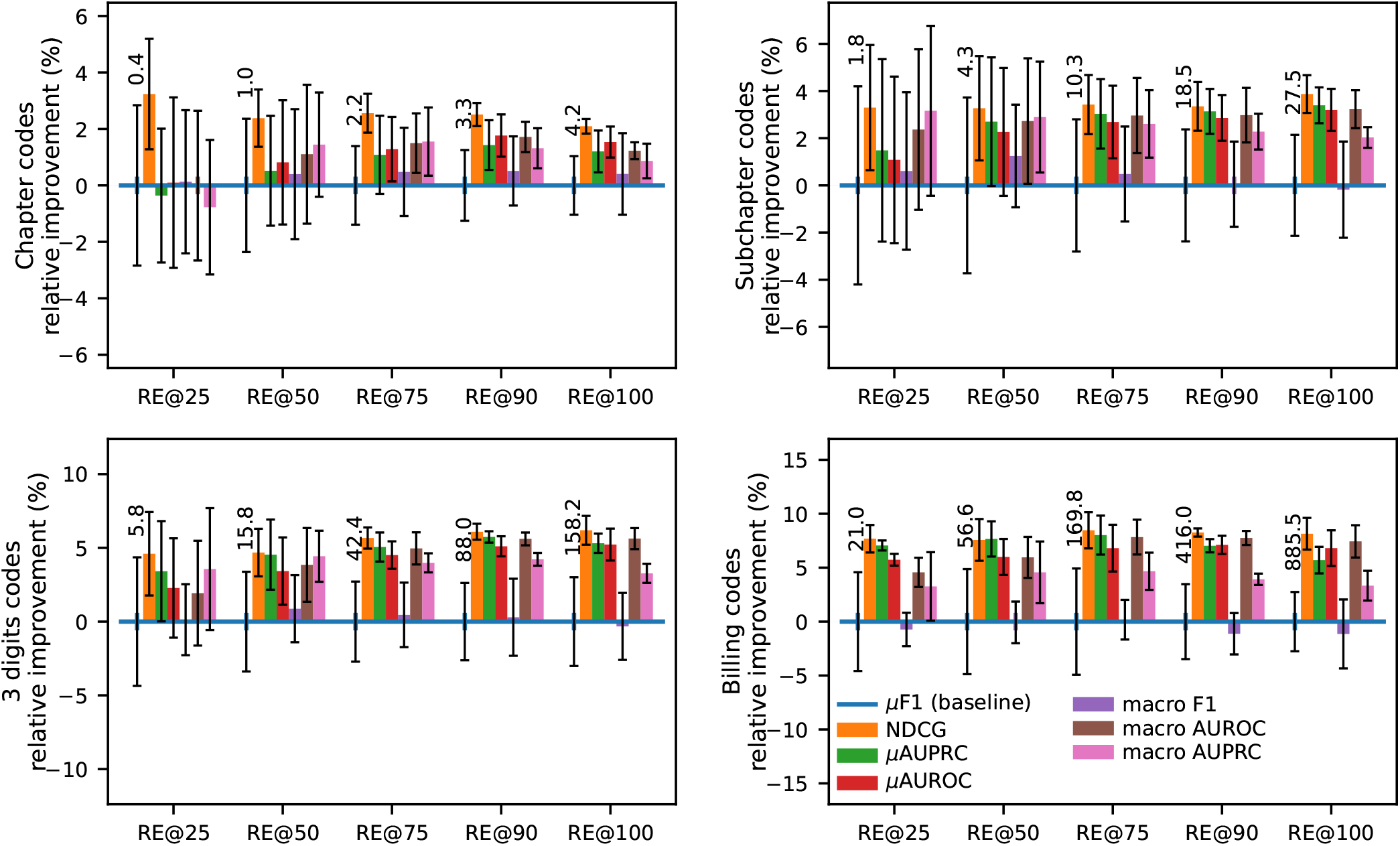
Relative improvement in ranking performance, measured by RE@R, for different hierarchy levels and various values of R achieved by optimizing various scalar performance metrics instead of the commonly reported *μ*F1. Error bars represent the standard error of the mean computed over cross-validation runs. We indicate, as reference, the absolute RE@R values, averaged over cross validation runs, achieved by the baseline model optimizing the *μ*F1.

**Fig. S4:**
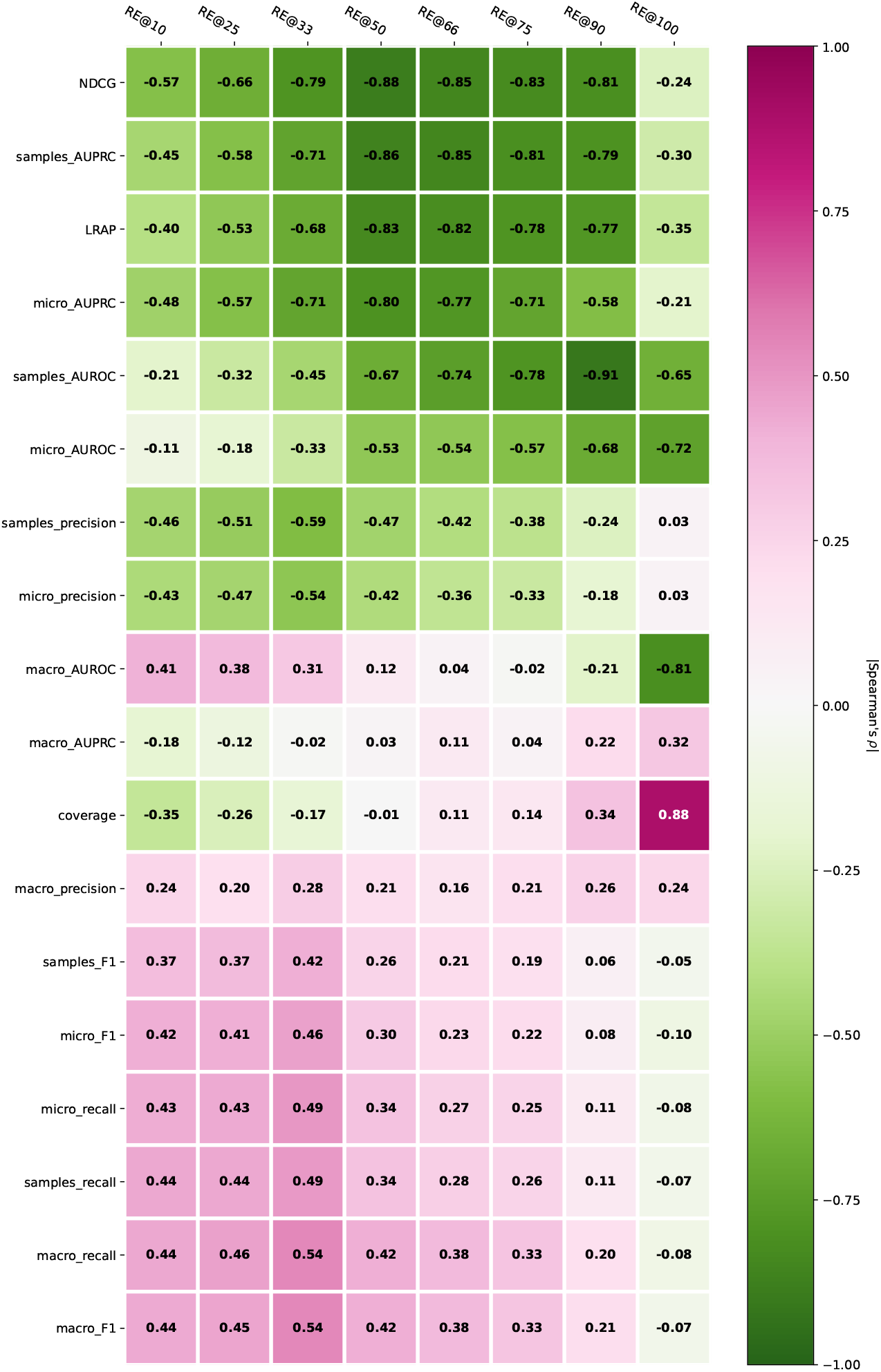
Spearman rank correlation between different metrics values and rank errors for different recall values evaluated on the complete code set and all code levels. Aiming to minimize ranking errors, the lower the correlation the better (except for coverage which we also aim to minimize). The metrics were sorted according to the average correlation value across RE@R for easier interpretation. To compute the different metrics we used the predictions from all 50 models trained on the complete code set presented in this paper and evaluated each metric on the complete code set.

**Fig. S5:**
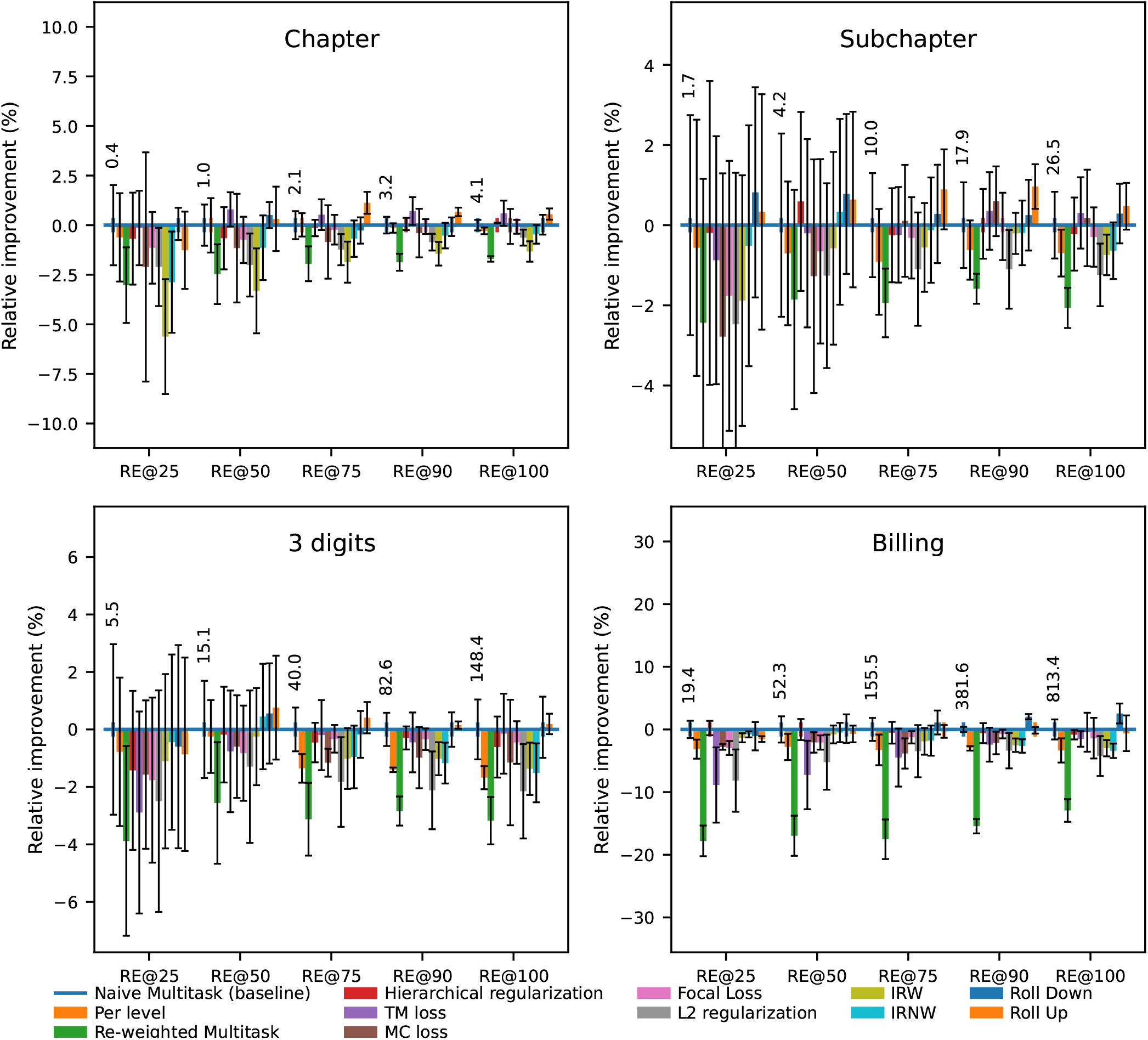
Relative improvement in ranking performance, quantified by RE@R, for different hierarchy levels and various values of R, achieved by models chosen to maximize global NDCG.

**Fig. S6:**
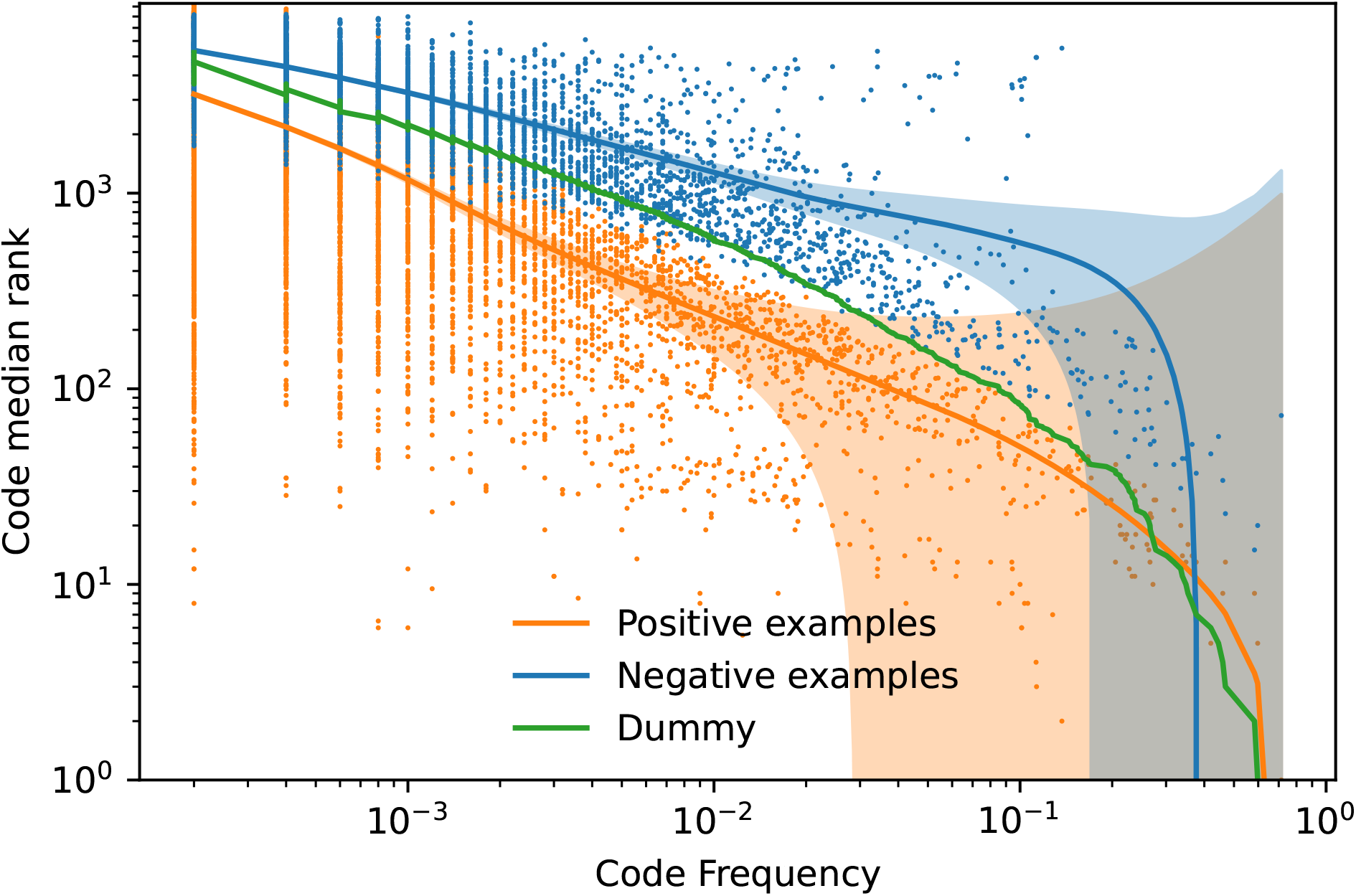
Relationship between ranks and code frequency. The lines and shaded areas were generated using LOWESS (Locally Weighted Scatterplot Smoothing) based on predictions from our top-performing NDCG-focused naive multitask model. The shaded regions represent the 95% confidence intervals. The presence of orange dots in the lower-left area demonstrates the algorithm’s capability to assign high ranks even to low-frequency codes.

**Fig. S7:**
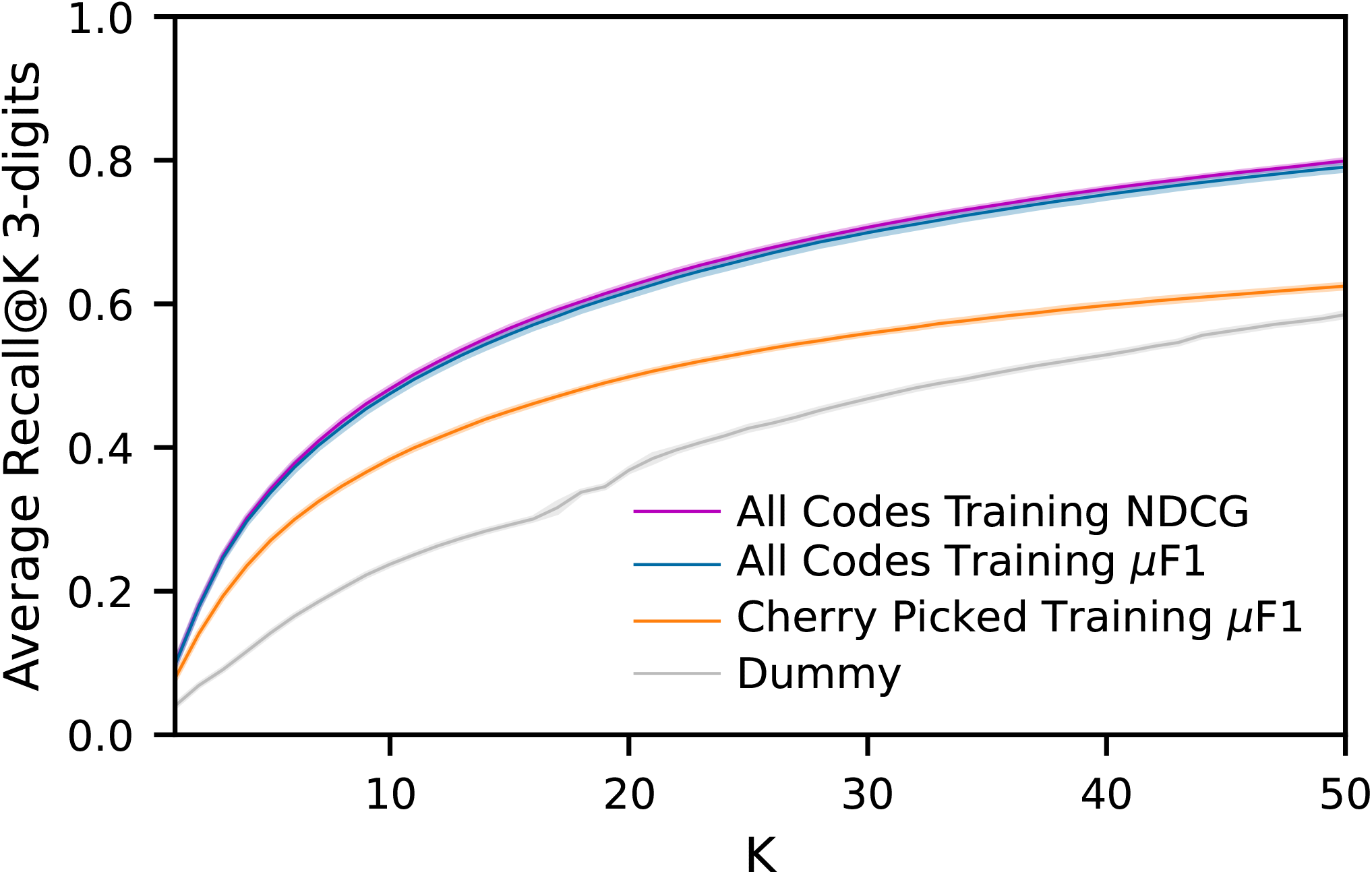
Comparison of our best model (purple) similar to the state of the art strategy in [S15](orange) for Recall@K on the 3-digits level.

## References

1. Rasmy, L., Xiang, Y., Xie, Z., Tao, C. & Zhi, D. Med-BERT: Pretrained Contextualized Embeddings on Large-Scale Structured Electronic Health Records for Disease Prediction. npj Digital Medicine 4, 1–13. ISSN: 2398-6352. (2023) (May 2021).

2. Stausberg, J., Lehmann, N., Kaczmarek, D. & Stein, M. Reliability of Diagnoses Coding with ICD-10. International Journal of Medical Informatics 77, 50–57. ISSN: 1386-5056 (Jan. 2008).

3. Johnson, A. E. W. et al. MIMIC-III, a Freely Accessible Critical Care Database. Scientific Data 3, 160035. ISSN: 2052-4463. (2021) (May 2016).

4. Farkas, R. & Szarvas, G. Automatic Construction of Rule-Based ICD-9-CM Coding Systems. BMC Bioinformatics 9, S10. ISSN: 1471-2105. (2022) (Apr. 2008).

5. Perotte, A. et al. Diagnosis Code Assignment: Models and Evaluation Metrics. Journal of the American Medical Informatics Association 21, 231–237. ISSN: 1067-5027, 1527-974X. (2021) (Mar. 2014).

6. Malakouti, S. & Hauskrecht, M. Not All Samples Are Equal: Class Dependent Hierarchical Multi-Task Learning for Patient Diagnosis Classification, 6.

7. Chalkidis, I. et al. An Empirical Study on Large-Scale Multi-Label Text Classification Including Few and Zero-Shot Labels. arXiv:2010.01653 [cs]. arXiv: 2010 . 01653 [cs]. (2021) (Oct. 2020).

8. Yogarajan, V., Montiel, J., Smith, T. & Pfahringer, B. Transformers for Multi-label Classification of Medical Text: An Empirical Comparison in Artificial Intelligence in Medicine (eds Tucker, A., Henriques Abreu, P., Cardoso, J., Pereira Rodrigues, P. & Riaño, D.) (Springer International Publishing, Cham, 2021), 114–123. ISBN: 978-3-030-77211-6.

9. Blinov, P., Avetisian, M., Kokh, V., Umerenkov, D. & Tuzhilin, A. Predicting Clinical Diagnosis from Patients Electronic Health Records Using BERT-Based Neural Networks in Artificial Intelligence in Medicine (eds Michalowski, M. & Moskovitch, R.) (Springer International Publishing, Cham, 2020), 111–121. ISBN: 978-3-030-59137-3.

10. Gao, S. et al. Limitations of Transformers on Clinical Text Classification. IEEE Journal of Biomedical and Health Informatics 25, 3596–3607. ISSN: 2168-2208 (Sept. 2021).

11. Xu, K. et al. Multimodal Machine Learning for Automated ICD Coding in Proceedings of the 4th Machine Learning for Healthcare Conference (PMLR, Oct. 2019), 197–215. (2022).

12. Li, F. & Yu, H. ICD Coding from Clinical Text Using Multi-Filter Residual Convolutional Neural Network Nov. 2019. arXiv: 1912 . 00862 [cs]. (2022).

13. Mullenbach, J., Wiegreffe, S., Duke, J., Sun, J. & Eisenstein, J. Explainable Prediction of Medical Codes from Clinical Text in Proceedings of the 2018 Conference of the North American Chapter of the Association for Computational Linguistics: Human Language Technologies, Volume 1 (Long Papers) (Association for Computational Linguistics, New Orleans, Louisiana, 2018), 1101–1111. (2021).

14. Rios, A. & Kavuluru, R. Few-Shot and Zero-Shot Multi-Label Learning for Structured Label Spaces in Proceedings of the 2018 Conference on Em-pirical Methods in Natural Language Processing (Association for Computational Linguistics, Brussels, Belgium, Oct. 2018), 3132–3142. (2022).

15. Teng, F., Yang, W., Chen, L., Huang, L. & Xu, Q. Explainable Prediction of Medical Codes With Knowledge Graphs. Frontiers in Bioengineering and Biotechnology 8, 867. ISSN: 2296-4185. (2021) (2020).

16. Catling, F., Spithourakis, G. P. & Riedel, S. Towards Automated Clinical Coding. International Journal of Medical Informatics 120, 50–61. ISSN: 13865056. (2022) (Dec. 2018).

17. Wang, S.-M. et al. Using Deep Learning for Automatic Icd-10 Classification from Free-Text Data. European Journal of Biomedical Informatics. (2021) (2020).

18. Sun, W., Ji, S., Cambria, E. & Marttinen, P. Multitask Recalibrated Aggregation Network for Medical Code Prediction in Machine Learning and Knowledge Discovery in Databases. Applied Data Science Track (eds Dong, Y., Kourtellis, N., Hammer, B. & Lozano, J. A.) (Springer Interna-tional Publishing, Cham, 2021), 367–383. ISBN: 978-3-030-86514-6.

19. Rodrigues-Jr, J., Gutierrez, M., Spadon, G., Brandoli, B. & Amer-Yahia, S. LIG-Doctor: Efficient Patient Trajectory Prediction Using Bidirectional Minimal Gated-Recurrent Networks. Information sciences, Information Sciences. (2021) (Feb. 2021).

20. Hansen, E. R. et al. Assigning Diagnosis Codes Using Medication History. Artificial Intelligence in Medicine 128, 102307. ISSN: 0933-3657. (2023) (June 2022).

21. Choi, E., Bahadori, M. T., Schuetz, A., Stewart, W. F. & Sun, J. Doctor AI: Predicting Clinical Events via Recurrent Neural Networks in Proceedings of the 1st Machine Learning for Healthcare Conference (PMLR, Dec. 2016), 301–318. (2022).

22. Zhou, L., Cheng, C., Ou, D. & Huang, H. Construction of a Semi-Automatic ICD-10 Coding System. BMC medical informatics and decision making 20, 67. ISSN: 1472-6947 (Apr. 2020).

23. Chen, P.-F. et al. Automatic ICD-10 Coding and Training System: Deep Neural Network Based on Supervised Learning. JMIR Medical Informatics 9, e23230. (2021) (Aug. 2021).

24. Campbell, S. & Giadresco, K. Computer-Assisted Clinical Coding: A Narrative Review of the Literature on Its Benefits, Limitations, Implementation and Impact on Clinical Coding Professionals. Health Information Management Journal 49, 5–18. ISSN: 1833-3583. (2022) (Jan. 2020).

25. Henry, K. E. et al. Human–Machine Teaming Is Key to AI Adoption: Clinicians’ Experiences with a Deployed Machine Learning System. npj Digital Medicine 5, 1–6. ISSN: 2398-6352. (2022) (July 2022).

26. Choi, E., Bahadori, M. T., Song, L., Stewart, W. F. & Sun, J. GRAM: Graph-based Attention Model for Healthcare Representation Learning in Pro-ceedings of the 23rd ACM SIGKDD International Conference on Knowledge Discovery and Data Mining (ACM, Halifax NS Canada, Aug. 2017), 787–795. ISBN: 978-1-4503-4887-4. (2022).

27. Cao, P. et al. HyperCore: Hyperbolic and Cograph Representation for Automatic ICD Coding in Proceedings of the 58th Annual Meeting of the Association for Computational Linguistics (Association for Computational Linguistics, Online, 2020), 3105–3114. (2022).

28. Malakouti, S. & Hauskrecht, M. Hierarchical Adaptive Multi-task Learning Framework for Patient Diagnoses and Diagnostic Category Classification in 2019 IEEE International Conference on Bioinformatics and Biomedicine (BIBM) (IEEE, San Diego, CA, USA, Nov. 2019), 701–706. ISBN: 978-1-72811-867-3. (2022).

29. Paris, N. & Parrot, A. MIMIC in the OMOP Common Data Model Aug. 2020. (2021).

30. Li, L., Jamieson, K., DeSalvo, G., Rostamizadeh, A. & Talwalkar, A. Hyperband: A Novel Bandit-Based Approach to Hyperparameter Optimization. Journal of Machine Learning Research 18, 1–52. ISSN: 1533-7928. (2023) (2018).

31. Wang, Y., Wang, L. & Li, Y. A Theoretical Analysis of Normalized Discounted Cumulative Gain (NDCG) Ranking Measures, 30.

32. Caruana, R. Multitask Learning, 35.

33. Li, L., Zhou, T., Wang, W., Li, J. & Yang, Y. Deep Hierarchical Semantic Segmentation. arXiv:2203.14335 [cs]. arXiv: 2203.14335 [cs]. (2022) (Mar. 2022).

34. Giunchiglia, E. & Lukasiewicz, T. Coherent Hierarchical Multi-Label Classification Networks, 12.

35. Gopal, S. & Yang, Y. Recursive Regularization for Large-Scale Classification with Hierarchical and Graphical Dependencies in Proceedings of the 19th ACM SIGKDD International Conference on Knowledge Discovery and Data Mining (ACM, Chicago Illinois USA, Aug. 2013), 257–265. ISBN: 978-1-4503-2174-7. (2021).

36. Naik, A. & Rangwala, H. Large Scale Hier-archical Classification: State of the Art ISBN: 978-3-030-01619-7 978-3-030-01620-3. (2021) (Springer International Publishing, Cham, 2018).

37. Guo, C., Pleiss, G., Sun, Y. & Weinberger, K. Q. On Calibration of Modern Neural Networks in Proceedings of the 34th International Conference on Machine Learning (PMLR, July 2017), 1321–1330. (2023).

## Supplementary References

S1. Wang, Y., Wang, L. & Li, Y. A Theoretical Analysis of Normalized Discounted Cumulative Gain (NDCG) Ranking Measures, 30.

S2. Wang, Y. et al. Clinical Information Extraction Applications: A Literature Review. Journal of Biomedical Informatics 77, 34–49. ISSN: 1532-0464. (2021) (Jan. 2018).

S3. Guo, C., Pleiss, G., Sun, Y. & Weinberger, K. Q. On Calibration of Modern Neural Networks in Proceedings of the 34th International Conference on Machine Learning (PMLR, July 2017), 1321–1330. (2023).

S4. Lin, T.-Y., Goyal, P., Girshick, R., He, K. & Dollár, P. Focal Loss for Dense Object Detection. arXiv:1708.02002 [cs]. arXiv: 1708.02002 [cs]. (2021) (Feb. 2018).

S5. Naik, A. & Rangwala, H. Large Scale Hierarchical Classification: State of the Art ISBN: 978-3-030-01619-7 978-3-030-01620-3. (2021) (Springer International Publishing, Cham,2018).

S6. Gopal, S. & Yang, Y. Recursive Regularization for Large-Scale Classification with Hierarchical and Graphical Dependencies in Proceedings of the 19th ACM SIGKDD International Conference on Knowledge Discovery and Data Mining (ACM, Chicago Illinois USA, Aug. 2013), 257–265. ISBN: 978-1-4503-2174-7. (2021).

S7. Charuvaka, A. & Rangwala, H. in Machine Learning and Knowledge Discovery in Databases (eds Appice, A. et al.) 675–690 (Springer International Publishing, Cham, 2015). ISBN: 978-3-319-23527-1 978-3-319-23528-8. (2021).

S8. Giunchiglia, E. & Lukasiewicz, T. Coherent Hierarchical Multi-Label Classification Networks, 12.

S9. Li, L., Zhou, T., Wang, W., Li, J. & Yang, Y. Deep Hierarchical Semantic Segmentation. arXiv:2203.14335 [cs]. arXiv: 2203.14335 [cs]. (2022) (Mar. 2022).

S10. Wehrmann, J., Cerri, R. & Barros, R. Hierarchical Multi-Label Classification Networks in Proceedings of the 35th International Conference on Machine Learning (PMLR, July 2018), 5075–5084. (2021).

S11. Fernández, A. et al. Learning from Imbalanced Data Sets ISBN: 978-3-319-98073-7 978-3-319-98074-4. (2021) (Springer International Publishing, Cham, 2018).

S12. Tarekegn, A. N., Giacobini, M. & Michalak, K. A Review of Methods for Imbalanced Multi-Label Classification. Pattern Recognition 118, 107965. ISSN: 00313203. (2021) (Oct. 2021).

S13. Pereira, R. M., Costa, Y. M. G. & Silla, C. N. Handling Imbalance in Hierarchical Classification Problems Using Local Classifiers Approaches. Data Mining and Knowledge Discovery 35, 1564–1621. ISSN: 1384-5810, 1573-756X. (2021) (July 2021).

S14. Miranda Pereira, R., Maldonado e Gomes da Costa, Y. & Nascimento Silla, C. Dealing with Imbalanceness in Hierarchical Multi-Label Datasets Using Multi-Label Resampling Techniques in 2018 IEEE 30th International Conference on Tools with Artificial Intelligence (ICTAI) (IEEE, Volos, Greece, Nov. 2018), 818–824. ISBN: 978-1-5386-7449-9. (2021).

S15. Hansen, E. R. et al. Assigning Diagnosis Codes Using Medication History. Artificial Intelligence in Medicine 128, 102307. ISSN: 0933-3657. (2023) (June 2022).

S16. Malakouti, S. & Hauskrecht, M. Hierarchical Adaptive Multi-task Learning Framework for Patient Diagnoses and Diagnostic Category Classification in 2019 IEEE International Conference on Bioinformatics and Biomedicine (BIBM) (IEEE, San Diego, CA, USA, Nov. 2019), 701–706. ISBN: 978-1-72811-867-3. (2022).

S17. Malakouti, S. & Hauskrecht, M. Not All Samples Are Equal: Class Dependent Hierarchical Multi-Task Learning for Patient Diagnosis Classification, 6.

S18. Mullenbach, J., Wiegreffe, S., Duke, J., Sun, J. & Eisenstein, J. Explainable Prediction of Medical Codes from Clinical Text in Proceedings of the 2018 Conference of the North American Chapter of the Association for Computational Linguistics: Human Language Technologies, Volume 1 (Long Papers) (Association for Computational Linguistics, New Orleans, Louisiana, 2018), 1101–1111. (2021).

S19. Yogarajan, V., Montiel, J., Smith, T. & Pfahringer, B. Transformers for Multi-label Classification of Medical Text: An Empirical Comparison in Artificial Intelligence in Medicine (eds Tucker, A., Henriques Abreu, P., Cardoso, J., Pereira Rodrigues, P. & Riaño, D.) (Springer International Publishing, Cham, 2021), 114–123. ISBN: 978-3-030-77211-6.

S20. Rodrigues-Jr, J., Gutierrez, M., Spadon, G., Brandoli, B. & Amer-Yahia, S. LIG-Doctor: Efficient Patient Trajectory Prediction Using Bidirectional Minimal Gated-Recurrent Networks. Information sciences, Information Sciences. (2021) (Feb. 2021).

S21. Perotte, A. et al. Diagnosis Code Assignment: Models and Evaluation Metrics. Journal of the American Medical Informatics Association 21, 231–237. ISSN: 1067-5027, 1527-974X. (2021) (Mar. 2014).

S22. Chalkidis, I. et al. An Empirical Study on Large-Scale Multi-Label Text Classification Including Few and Zero-Shot Labels. arXiv:2010.01653 [cs]. arXiv: 2010 . 01653 [cs]. (2021) (Oct. 2020).

S23. Gao, S. et al. Limitations of Transformers on Clinical Text Classification. IEEE Journal of Biomedical and Health Informatics 25, 3596–3607. ISSN: 2168-2208 (Sept. 2021).

S24. Sonabend W A. et al. Automated ICD Coding via Unsupervised Knowledge Integration (UNITE). International Journal of Medical Informatics 139, 104135. ISSN: 13865056. (2022) (July 2020).

S25. Li, M. et al. Automated ICD-9 Coding via A Deep Learning Approach. IEEE/ACM Transactions on Computational Biology and Bioinformatics 16, 1193–1202. ISSN: 1557-9964 (July 2019).

S26. Wanyan, T. et al. Heterogeneous Graph Embeddings of Electronic Health Records Improve Critical Care Disease Predictions in Artificial Intelligence in Medicine (eds Michalowski, M. & Moskovitch, R.) (Springer International Publishing, Cham, 2020), 14–25. ISBN: 978-3-030-59137-3.

S27. Sun, W., Ji, S., Cambria, E. & Marttinen, P. Multitask Recalibrated Aggregation Network for Medical Code Prediction in Machine Learning and Knowledge Discovery in Databases. Applied Data Science Track (eds Dong, Y., Kourtellis, N., Hammer, B. & Lozano, J. A.) (Springer International Publishing, Cham, 2021), 367–383. ISBN: 978-3-030-86514-6.

S28. Catling, F., Spithourakis, G. P. & Riedel, S. Towards Automated Clinical Coding. International Journal of Medical Informatics 120, 50–61. ISSN: 13865056. (2022) (Dec. 2018).

S29. Xu, K. et al. Multimodal Machine Learning for Automated ICD Coding in Proceedings of the 4th Machine Learning for Healthcare Conference (PMLR, Oct. 2019), 197–215. (2022).

S30. Cao, P. et al. HyperCore: Hyperbolic and Cograph Representation for Automatic ICD Coding in Proceedings of the 58th Annual Meeting of the Association for Computational Linguistics (Association for Computational Linguistics, Online, 2020), 3105–3114. (2022).

S31. Li, F. & Yu, H. ICD Coding from Clinical Text Using Multi-Filter Residual Convolutional Neural Network Nov. 2019. arXiv: 1912 . 00862 [cs]. (2022).

